# Multivariate, Multi-omic Analysis in 799,429 Individuals Identifies 134 Loci Associated with Somatoform Traits

**DOI:** 10.1101/2024.07.29.24310991

**Authors:** Christal N. Davis, Sylvanus Toikumo, Alexander S. Hatoum, Yousef Khan, Benjamin K. Pham, Shreya R. Pakala, Kyra L. Feuer, Joel Gelernter, Sandra Sanchez-Roige, Rachel L. Kember, Henry R. Kranzler

## Abstract

Somatoform traits, which manifest as persistent physical symptoms without a clear medical cause, are prevalent and pose challenges to clinical practice. Understanding the genetic basis of these disorders could improve diagnostic and therapeutic approaches. With publicly available summary statistics, we conducted a multivariate genome-wide association study (GWAS) and multi-omic analysis of four somatoform traits—fatigue, irritable bowel syndrome, pain intensity, and health satisfaction—in 799,429 individuals genetically similar to Europeans. Using genomic structural equation modeling, GWAS identified 134 loci significantly associated with a somatoform common factor, including 44 loci not significant in the input GWAS and 8 novel loci for somatoform traits. Gene-property analyses highlighted an enrichment of genes involved in synaptic transmission and enriched gene expression in 12 brain tissues. Six genes, including members of the CD300 family, had putatively causal effects mediated by protein abundance. There was substantial polygenic overlap (76-83%) between the somatoform and externalizing, internalizing, and general psychopathology factors. Somatoform polygenic scores were associated most strongly with obesity, Type 2 diabetes, tobacco use disorder, and mood/anxiety disorders in independent biobanks. Drug repurposing analyses suggested potential therapeutic targets, including MEK inhibitors. Mendelian randomization indicated potentially protective effects of gut microbiota, including *Ruminococcus bromii*. These biological insights provide promising avenues for treatment development.

## Introduction

Persistent physical symptoms (PPS) adversely impact the quality of life of affected individuals, increase their healthcare utilization, and contribute to disability as much as or more than etiologically well-defined medical diseases.^1^ PPS have historically been referred to as medically unexplained symptoms, but this terminology fails to reflect the dynamic nature of medical knowledge and contributes to a false contrast between these symptoms and “real” medical diseases.^2^ PPS may occur alone or as part of a functional somatic syndrome (FSS), such as irritable bowel syndrome (IBS), fibromyalgia, or myalgic encephalomyelitis/chronic fatigue syndrome (ME/CFS). It is estimated that up to 10% of the general population is affected by at least one FSS,^3^ with much higher rates likely at the symptom level. In a meta-analysis of over 70,000 primary care patients from 24 countries, ∼45% reported at least one somatic symptom with no identified organic cause.^4^ Symptoms that general practitioners deem medically unexplained occur at higher rates among women and non-native speakers,^5^ potentially reflecting biases in diagnostic practices and healthcare delivery. Despite their prevalence and impact, the underlying mechanisms of PPS remain poorly understood.

The various presentations of PPS have complex genetic and environmental etiologies, further complicated by frequently co-occurring psychiatric disorders.^1,6^ Genome-wide association studies (GWAS), which aim to identify genetic risk variants for complex traits and diseases, have revealed loci associated with IBS,^7^ chronic pain,^8^ subjective perceptions of pain intensity,^9^ and headaches/migraine.^10^ These GWAS implicate gene expression and biological processes in the immune and central nervous systems as having key roles in the etiology of PPS. Furthermore, significant genetic correlations exist between different PPS *and* between PPS and autoimmune, psychiatric, and anthropometric traits.^9,11,12^ The substantial phenotypic and genetic correlations between different PPS and between PPS and psychiatric conditions suggest a shared etiology.^13,14^ Aligned with this perspective, the Hierarchical Taxonomy of Psychopathology (HiTOP)—an empirical nosology model—proposes that a somatoform spectrum reflects the common liability to PPS.^15,16^ Furthermore, reflecting the commonality of somatoform traits with psychiatric conditions, the HiTOP somatoform spectrum is subsumed under a broader emotional dysfunction superspectrum.^16^

Previous work using genomic structural equation modeling (gSEM) provided evidence that chronic pain conditions have a shared genetic basis.^12^ Other studies demonstrated shared genetic variation among six nociplastic pain conditions (i.e., those in which there is persistent pain without tissue damage): chronic widespread pain (CWP), endometriosis, low back pain, broadly defined headache, irritable bowel syndrome (IBS), and temporomandibular joint disorder (TMJD).^11^ However, this past work focused largely on pain rather than the broader somatoform spectrum, and some of the included GWAS had modest sample sizes (e.g., TMJD: N_cases_ = 217, CWP: N_cases_ = 6,914), which may have limited the strength and precision of the identified common factor. A deeper understanding of the shared genetic basis across a broader spectrum of PPS could identify contributory factors and biological pathways that underpin these syndromes. Genetic research on somatoform traits and their potential polygenic overlap with psychiatric conditions could also help to refine the current diagnostic system, improving the assessment and treatment of PPS.

Because our understanding of the genetic basis of the broader PPS spectrum remains limited, we leveraged gSEM to examine the shared genetic architecture of fatigue, health satisfaction, IBS, and pain intensity. Although we initially included headache/migraine as well, it was not retained due to poorer fit on the latent factor. These traits index a possible somatoform spectrum, as they encompass both PPS and symptom perceptions.^16^ Next, we performed a comprehensive set of bioinformatic analyses using the GWAS summary statistics (Figure 1). Our gene prioritization efforts included gene mapping, functional annotation, transcriptome-wide association studies (TWAS) in enriched tissues, and the integration of brain eQTL and blood plasma pQTL data using summary-data-based Mendelian randomization (SMR) to identify putative causal genes. To characterize the genetic architecture of the somatoform factor more broadly, we performed MAGMA gene-property analyses, univariate and bivariate causal mixture models (MiXeR), batch genetic correlations with 1,426 publicly available GWAS, and phenome-wide association studies (PheWAS) in three cohorts (i.e., BioVU, Penn Medicine BioBank, and Yale-Penn). Finally, we conducted two sets of analyses aimed at potential translational applications of GWAS—drug repurposing and Mendelian randomization to identify causal effects of the gut microbiome on the somatoform factor given emerging evidence of the role of the gut-brain-axis in regulating pain.^17^ With this approach, we aimed to deepen our understanding of the genetic basis of somatoform traits, contribute to the refinement of existing diagnostic models, and identify potential treatments.

**Figure 1.**
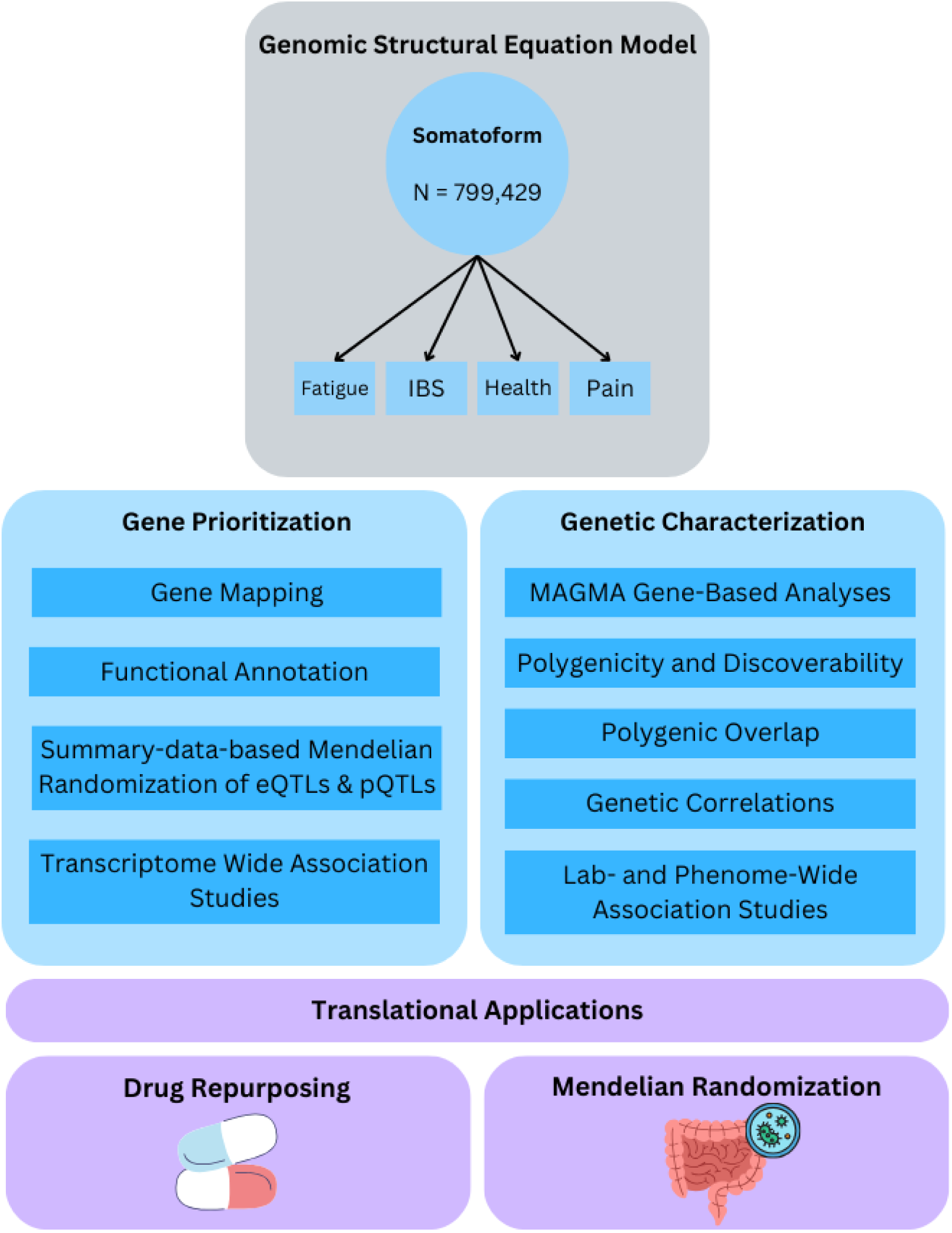
Overview of analyses. *Note:* IBS = irritable bowel syndrome. The somatoform factor N reflects the effective sample size.

## Methods

### Summary Statistics

Summary statistics were chosen to correspond to the HiTOP somatoform factor,^15^ which consists of conversion (i.e., neurological symptoms that lack an identified medical cause), somatization (i.e., the tendency to express psychological distress in the form of physical symptoms), malaise (i.e., a general sense of poor wellbeing), head pain, gastrointestinal, and cognitive (e.g., illness anxiety) symptoms. We acknowledge the historical stigma associated with terms like “conversion” and “somatization”^18^ and use them here solely to establish consistency with the HiTOP model. In line with patient advocacy groups, we encourage the use of alternate terms like FSS and PPS.

We selected five sets of publicly available GWAS summary statistics from well-powered studies of individuals genetically similar to Europeans (EUR):^19^ fatigue (N = 350,580; http://www.nealelab.is/uk-biobank/), headache and migraine^10^ (N = 360,391; ultimately not retained due to poorer common factor fit), health satisfaction (N = 119,567; http://www.nealelab.is/uk-biobank/), IBS^7^ (N = 486,601), and pain intensity^9^ (N = 436,683). The input summary statistics included between 6,788,440 (pain intensity) and 13,586,245 (fatigue) SNPs. Health satisfaction was reverse coded so that higher scores indicated lower satisfaction to maintain consistency of risk direction with the other traits.

### Genomic Structural Equation Modeling

Using the *GenomicSEM* R package,^20^ standard quality controls were applied to the input summary statistics, including filtering to EUR HapMap3 SNPs,^21^ and when the information was available, retaining only SNPs with MAF > 0.01 and INFO > 0.6. Linkage disequilibrium score regression (LDSC) was implemented within *GenomicSEM* to estimate genetic covariance matrices for the input traits. The resulting LDSC matrices were then used to perform confirmatory factor analysis (CFA) to determine whether the hypothesized common factor model fit the data well. Model fit was evaluated based on chi-square (non-significance indicates better fit), Akaike information criterion (AIC; lower values indicate better relative fit), comparative fit index (CFI; > 0.9 indicates good fit), and standardized root mean square residual (SRMR; < 0.08 indicates good fit) values. We also examined the proportion of variance in each input trait that was explained by the common factor to ensure that each trait was sufficiently represented by the factor, aiming for standardized loadings of at least 0.30.

To perform common factor GWAS, we regressed each SNP on the somatoform latent variable. Q_SNP_ was used to identify any SNPs that had heterogeneous effects across the input traits. SNPs with a significant Q_SNP_ value (*p* < 5e-8) were removed from the summary statistics and not included in any subsequent analyses, as these SNPs’ effects were not well represented by a common factor. The effective sample size of the resulting GWAS was calculated using the formula described by Mallard et al. (2022).^22^ Clumping of the GWAS results was performed using PLINK 1.9^23^ with an *r*^2^ threshold of 0.1 and a physical distance threshold of 3000 kb. The novelty of loci was based on the lead SNP’s positional overlap (within ± 1000 kb) with genome-wide significant (GWS) variants from previous GWAS of the included traits. For all novel SNPs, we performed SNP-level PheWAS using both GWAS Atlas^24^ and the *ieugwasr* R package from the OpenGWAS database.^25^

### Gene Mapping and Functional Annotation

Using the SNP2GENE function in FUMA v1.5.2^26^, SNPs were mapped to protein-coding genes using three approaches: (1) position (≤ 10kb), (2) eQTL (BrainSeq,^27^ PsychENCODE,^28^ BRAINEAC,^29^ and GTEx v8^30^ brain tissue data), and (3) chromatin interaction mapping (Hi-C brain tissues).^31,32^ SNPs were functionally annotated using ANNOVAR,^33^ Combined Annotation Dependent Depletion (CADD),^34^ RegulomeDB,^35^ and PsychENCODE^28^ databases. ANNOVAR identifies the functional consequences of SNPs, such as whether they fall within exons, introns, or regulatory regions. Functional predictions also included protein-coding changes (synonymous, nonsynonymous, stop-gain/loss) and splicing effects. CADD scores were used to assess the deleteriousness of SNPs, with higher scores indicating a greater likelihood of pathogenicity. We considered SNPs with CADD scores above 20 to be potentially deleterious and those above 12.37 (top 7.5%) to be likely pathogenic.^36^ RegulomeDB v2.2 was used to predict the regulatory potential of SNPs using functional data from eQTL, chromatin states, transcription factor binding sites, and other regulatory elements.^37^ Lower RegulomeDB scores indicate higher confidence in regulatory function. PsychENCODE data provided additional context on the regulatory role of SNPs in brain tissues.

### MAGMA Gene-Based Analyses

We performed gene-set and gene-property analyses using MAGMA v1.08.^38^ First, SNPs were positionally mapped (0 kb window) to protein-coding genes. Using the resulting gene-level *p*-values, gene-set analyses were performed for MsigDB v7.0^39^ curated gene sets and gene ontology (GO) terms. Gene-property analyses were performed for 54 tissue types (GTEx v8)^30^ and 11 developmental stages in brain samples (BrainSpan).^32^ These analyses test whether genes having certain properties (i.e., expression in a particular tissue or developmental period) are more likely to be associated with the somatoform factor, adjusting for the average expression of all tissues/developmental stages in the dataset. Additionally, cell-type-specificity analyses were conducted across 16 human brain cell datasets to investigate whether specific cell types were implicated in somatoform traits.

### eQTL and pQTL Association Analyses

To identify SNPs with associations to somatoform traits mediated by effects on gene and protein expression, we performed summary-data-based Mendelian randomization (SMR) analyses.^40^ The heterogeneity in dependent instruments (HEIDI) test was used to distinguish pleiotropic effects from linkage between somatoform traits and eQTL/pQTLs. We identified significant associations as those that had a *P*_SMR_ < 0.05 after Bonferroni correction for the number of genes tested and a *P*_HEIDI_ > 0.05. We used the MetaBrain^41^ *cis*-eQTL database from 7 brain regions due to its large sample size (*n* = 8,613) and comprehensive integration of data across 14 cohorts. For the blood pQTL analyses, we used two *cis-* and *trans*-pQTL databases. The first was the UK Biobank Pharma Proteomics Project (UK-PPP) that included samples from 54,219 individuals and measured 2,923 unique proteins.^42^ The second database was from the deCODE Consortium, including samples from 35,559 individuals and 4,719 unique proteins.^43^ Use of the two databases allowed for validation and replication of findings and leveraged varied proteomic approaches for more comprehensive protein coverage.

### Transcriptome-Wide Association Studies

We conducted two transcriptome-wide association studies (TWAS) using MetaXcan software.^44,45^ In the first, we used S-MultiXcan to simultaneously examine associations across all GTEx v8 tissues for which gene expression was enriched based on MAGMA results. S-MultiXcan accounts for the correlation between gene expression profiles across different tissues, increasing statistical power to detect biologically meaningful associations.^45^ We complemented this approach with S-PrediXcan analyses using data from PsychENCODE (i.e., 2,188 postmortem frontal and temporal cerebral cortex samples from 1,695 adults),^46^ which is enriched for individuals diagnosed with psychiatric conditions that are often comorbid with functional somatic conditions.^6^ This provides a more clinically relevant cohort for studying transcriptomic associations.

### Polygenic Overlap with Psychopathology

MiXeR software^53,54^ was used to conduct univariate and bivariate causal mixture models. Univariate causal mixture models estimate the polygenicity (i.e., the number of causal variants needed to explain 90% SNP-heritability) and discoverability (i.e., the average effect size of causal variants) of traits. Bivariate causal mixture models can estimate genetic overlap between traits, even when the causal variants have opposite directions of effect on the traits. The Dice coefficient estimates the proportion of polygenic SNPs out of the total number of estimated causal SNPs for both traits. We performed bivariate MiXeR analyses to estimate the degree of polygenic overlap between the somatoform factor, externalizing, internalizing, and general psychopathology factors.^55^ We selected these factors because of their inclusion in the HiTOP model^56^ and the high rates of comorbidity between somatoform traits and psychopathology.^3^

### Genetic Correlations with Publicly Available GWAS

We performed batch genetic correlations with 1,426 phenotypes from publicly available GWAS using the Complex-Trait Genetics Virtual Lab (CTG-VL).^47^ The included phenotypes spanned a variety of domains, including biological variables, physical diseases, and psychiatric disorders. CTG-VL uses LDSC software^48,49^ with 1000 Genomes Project phase 3^50^ EUR data as LD references. We applied a Bonferroni correction (*p* = 0.05/1426 = 3.51e-5) to account for multiple testing and identify significant correlations.

### Lab-and Phenome-Wide Association Studies

We conducted phenome-wide association studies (PheWAS) using somatoform polygenic scores (PGS) in three cohorts: BioVU, Penn Medicine BioBank (PMBB), and Yale-Penn. ICD-9 and ICD-10 codes from electronic health records (EHRs) or diagnostic interviews were mapped to phecodes. Lab-wide association studies (LabWAS) were also performed in BioVU to examine associations with lab test results and biomarkers.^51^ We calculated PGS using PRS-CS software,^52^ applying the default settings to estimate shrinkage parameters. PheWAS analyses were conducted in the *PheWAS* v0.12 R package using logistic or linear regression models, depending on the phenotype. Analyses were restricted to phenotypes that had at least 100 cases (for binary traits) or individuals assessed (for continuous traits). All models included age, sex, and the first ten genetic ancestry principal components (PCs) as covariates, with a Bonferroni correction applied to identify significant associations. Given that both BioVU and PMBB are EHR datasets, we meta-analyzed the PheWAS results from the two cohorts. For each unique phenotype (1,442 total), we pooled the effect sizes by calculating a weighted average using the beta estimates and standard errors from each cohort.

*BioVU.* The BioVU cohort comprises Vanderbilt University Medical Center patients with EHR and genotype data.^53^ As described elsewhere,^51^ genotyping was performed using the Illumina Multi-Ethnic Genotype Array (MEGAEX). Genotypes were filtered for SNP (< 0.95) and individual (< 0.98) call rates, sex discrepancies, and excessive heterozygosity (|F_het_| > 0.2).^54^ PCA was performed using *FlashPCA2* 1000 Genomes phase 3 reference datasets^50^ to identify European-like individuals. Genotypes were imputed using the Michigan Imputation Server (https://imputationserver.sph.umich.edu) with the Haplotype Reference Consortium panel.^55^ SNPs with imputation quality R2 > 0.3 or INFO > 0.95 and MAF > 0.01 were retained. PheWAS analyses were performed in the EUR cohort (up to 66,214 individuals) on 1,338 case/control phecodes. To identify associations with biomarkers, we performed LabWAS for 315 biomarkers using a pipeline developed by Dennis et al.^51^

*Penn Medicine BioBank.* The Penn Medicine BioBank (PMBB) comprises a cohort recruited through the University of Pennsylvania Health System.^56^ Genotyping was performed using the GSA array, phasing was performed using EAGLE,^57^ and imputation was performed using Minimac4^58^ on the TOPMed Imputation server.^58^ Variants with imputation quality □<□0.7, missingness□>□5%, MAF□<□1%, and sample call rate□<□0.99 were excluded. PCs for genetic ancestry were calculated in EIGENSOFT v7.2.0 (https://github.com/DReichLab/EIG). Genetically inferred ancestry was assigned based on the distance of ten PCs from the 1000 Genomes^50^ reference populations. We performed PheWAS using 920 phenotypes in up to 29,090 EUR individuals.

*Yale-Penn.* The Yale-Penn sample, enriched for individuals with substance use disorders (SUDs), includes 5,424 EUR individuals with genotype data.^59^ As described previously,^59^ genotyping was performed using the Illumina HumanOmni1-Quad microarray, the Illumina HumanCoreExome array, or the Illumina Multi-Ethnic Global array. Imputation was performed using Minimac3^58^ and the 1000 Genomes Project phase 3^50^ reference panel on the Michigan Imputation Server (https://imputationserver.sph.umich.edu). SNPs with imputation quality < 0.7, MAF < 0.01, missingness > 0.01, or a batch allele frequency difference > 0.04 were excluded, as were individuals with genotype call rate < 0.95.^59^ PCs were used to determine genetic similarity to 1000 Genomes Project phase 3^50^ reference genomes. Yale-Penn participants were interviewed with the Semi-Structured Assessment for Drug Dependence and Alcoholism (SSADDA).^60^ PheWAS analyses were performed for 622 traits.

### Drug Repurposing

To perform drug repurposing, we used the Library of Integrated Network-Based Cellular Signatures (LINCS) L1000 database, which catalogs *in vitro* gene expression profiles from thousands of chemical compounds across more than 80 human cell lines. We focused on compounds that are either approved by the Food and Drug Administration (FDA) or currently undergoing clinical trials (https://clue.io/repurposing#download-data),^61^ yielding a total of 829 compounds (590 of which are FDA-approved) and expression profiles from five neuronal cell lines. This resulted in 3,897 unique signatures. We then matched medication signatures to gene expression signatures obtained from a somatoform TWAS performed on 12 enriched brain tissues. Weighted Pearson correlations were calculated between each brain transcriptome association and the compound signatures,^62^ with genes weighted by the proportion of heritability explained using the metafor *R* package (v.3.8-1). Each compound was included as a fixed effect, incorporating the weighted effect size (r_weighted) and sampling variability (se2_r_weighted) from all its signatures across different times, cell lines, and doses. To account for potential heterogeneity across tissues, brain region was included as a random effect. Significance was evaluated using a Bonferroni correction (*p* = 0.05/829 = 6.03e-5).

We also used a second genetically informed drug repurposing method, drug-gene set analysis (DRUGSETS),^63^ with data sourced from the Drug Repurposing Hub^61^ and the Drug Gene Interaction Database.^64^ For this method, drug–gene sets were created for 1,201 drugs, consisting of genes whose protein products are targeted by or interact with each drug. Competitive gene-set analysis was conducted using MAGMA v.1.08,^38^ conditioning on a comprehensive set of all drug targeted genes in the data (n = 2,116 across 735 gene sets). To determine significant drug-gene sets, we applied a Bonferroni correction (*p* = 0.05/735 = 6.80e-5).

### Mendelian Randomization with Gut Microbiome Taxa

To identify potential causal effects of the gut microbiome on somatoform traits, we conducted Mendelian randomization (MR) analyses using *TwoSampleMR* version 0.5.10 in R.^65^ We selected two GWAS of gut microbiota abundance to examine in relation to somatoform traits based on their complementary strengths. The first included 211 taxa (with genus being the most granular level) measured in 14,306 EUR individuals.^66^ The second included 207 taxa, including species level information, measured in a smaller sample of 7,738 Dutch individuals.^67^ Through the IEU GWAS database, we extracted instruments for each of the taxa at a *p*-value threshold of 1e-5. Instruments were clumped using EUR 1000 Genomes Project data to ensure independence. Matching SNPs were then extracted from the somatoform GWAS for use as outcomes. If an instrument SNP was not available in the somatoform GWAS, we selected proxy SNPs using the default settings in *TwoSampleMR*. The inverse variance weighted estimate was the primary MR method used for inferring causal effects. As sensitivity analyses, we performed tests of heterogeneity and horizontal pleiotropy, which assess whether the causal effects are consistent across all instruments and whether the instruments affect the outcome through pathways other than the exposure. We also performed the Steiger test, which can be used to evaluate whether the specified causal direction (i.e., microbiome → somatoform) is likely to be correct or if effects may operate in the opposite direction.^68^

## Results

Fatigue, headache/migraine, health satisfaction, IBS, and pain intensity all had significant SNP-heritability, ranging from 0.059 (SE = 0.003) for fatigue to 0.086 (SE = 0.004) for pain. LDSC identified significant genetic correlations between all the included traits (Supplementary Figure 1). However, genetic correlations with headache/migraine were lower than the other traits (*rg*_average_ = 0.27 vs. 0.58). A common factor CFA fit the data well (*χ*^2^(5) = 21.61, *p* = 0.001, AIC = 41.61, CFI = 0.99, SRMR = 0.03; Figure 2b). Standardized loadings ranged from 0.35 (for headache/migraine) to 0.89 (for fatigue). Although the loading of headache/migraine onto the common factor met our pre-specified minimum of 0.30, it was substantially lower than the other traits (all >0.65), suggesting that headache/migraine was not adequately represented by the common factor. To maintain a higher standard of representation and consistency across the included traits, we excluded headache/migraine from the final common factor model. After its exclusion, model fit improved significantly (Δ*χ*^2^(3) = 12.31, *p* = 0.006; fit statistics:*χ*^2^(2) = 9.30, *p* = 0.01, AIC = 25.30, CFI = 1, SRMR = 0.03).

**Figure 2.**
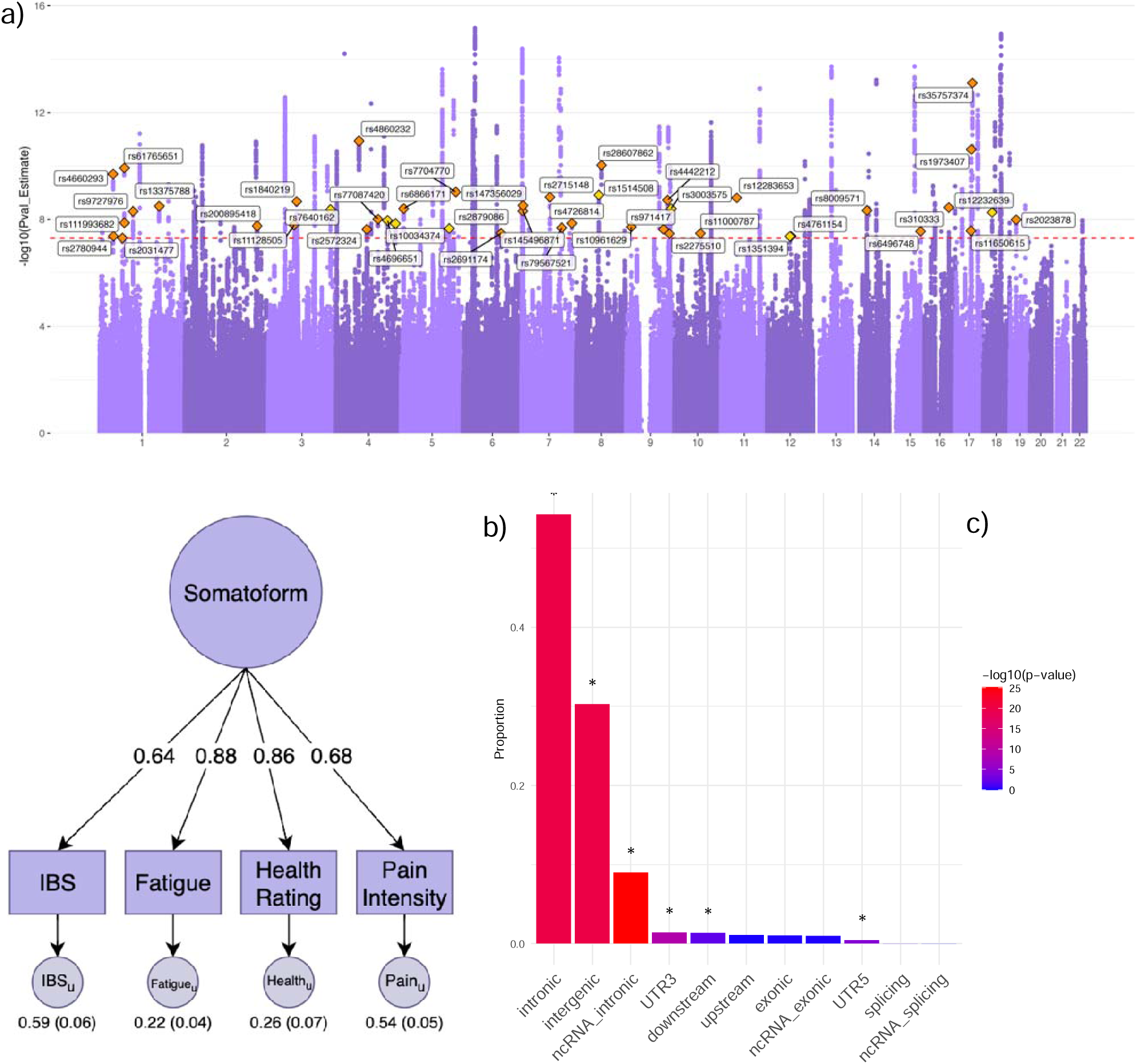
GWAS results for the somatoform factor. *Note:* **a)** Manhattan plot of the somatoform GWAS. Lead SNPs for loci not identified in the input GWAS are annotated. Gold diamonds indicate that the SNP was not in a genome-wide significant (GWS) locus in any of the input GWAS, and yellow diamonds indicate that the SNP was not in a GWS locus in previous GWAS of somatoform traits based on a GWAS Catalog search, **b)** confirmatory factor analysis, and **c)** functional consequences of SNPs on genes.

GWAS of the somatoform factor identified 134 significantly associated loci (N_eff_ = 799,429; Supplementary Table 1), of which 44 were not GWS in any of the input GWAS, and 8 had not been previously associated with somatoform traits (Figure 2a). The most strongly associated locus (*p* = 6.80e-16) was just outside the MHC region of chromosome 6, with the nearest gene being *UHRF1BP1*. *UHRF1BP1* is involved in assisting or regulating epigenetic modifications, particularly DNA methylation and chromatin remodeling.^69^ The lead SNP in this locus (rs9469907) functions as an eQTL for several genes, including *C6orf106*, *SNRPC*, and *CLPS*, in PsychENCODE and GTEx brain tissues.

In SNP-level PheWAS (Supplementary Table 2 and Supplementary Figure 2), the lead SNPs from the 8 novel loci were associated primarily with physical health, immunological, and mental health measures (e.g., insomnia, C-reactive protein levels, body mass index, wellbeing, depression, and Type 2 diabetes). Based on the *Q*_SNP_ test, 8 loci exhibited heterogeneous effects across the four somatoform traits (Supplementary Table 3). Of the lead SNPs from these loci, all but one showed the strongest association with pain intensity. The exception, rs10795616, was most strongly associated with health satisfaction (*p* = 2.21e-08).

### Gene Mapping and Functional Annotation

MAGMA identified 874 genes based on position, eQTL, and chromatin interactions. One-third (33.98%; n = 297) of genes were mapped by more than one approach, and 11.21% (n = 98) were mapped by all three. eQTL and chromatin interaction plots are shown in Supplementary Figure 3. Candidate SNPs showed an enrichment of intronic (*p* = 7.95e-317), intergenic (*p* = 7.43e-297), non-coding RNA intronic (*p* = 1.08e-8), 3’ UTR (*p* = 7.87e-7), and 5’ UTR (*p* = 2.62e-4) functional categories using ANNOVAR annotations (Figure 2c). Of candidate SNPs (n = 5,555), 30 had a CADD score > 20, and 255 (4.59%) had scores ≥12.37, which is suggestive of deleteriousness to gene function (Supplementary Table 4). Of the candidate SNPs, 59.17% (n = 3287) had RegulomeDB scores indicative of regulatory functions related to transcription factor binding and gene expression (i.e., 1a-1f; Supplementary Table 5).

### Gene-Based Enrichment

MAGMA gene-set analyses showed enrichment for genes involved in the biological process of negative regulation of synaptic transmission (b = 0.71, SE = 0.14, *p* = 3.24e-07). Gene-property analyses identified significant gene enrichment in 11 of 13 GTEx v8 brain tissues (Figure 3a). The strongest associations were for the cerebellar hemisphere (b = 0.05, SE = 0.01, *p* = 1.43e-12), cerebellum (b = 0.05, SE = 0.01, *p* = 2.63e-12), and frontal cortex (b = 0.05, SE = 0.01, *p* = 1.97e-9). In addition, there was significant enrichment for gene expression in the pituitary gland (b = 0.05, SE = 0.01, *p* = 5.64e-6). Gene-property analyses using BrainSpan brain samples from 11 developmental stages showed enrichment for gene expression in the early mid-prenatal (b = 0.04, SE = 0.01, *p* = 0.001) and the late mid-prenatal (b = 0.05, SE = 0.02, *p* = 0.002) stages (Supplementary Figure 4).

**Figure 3.**
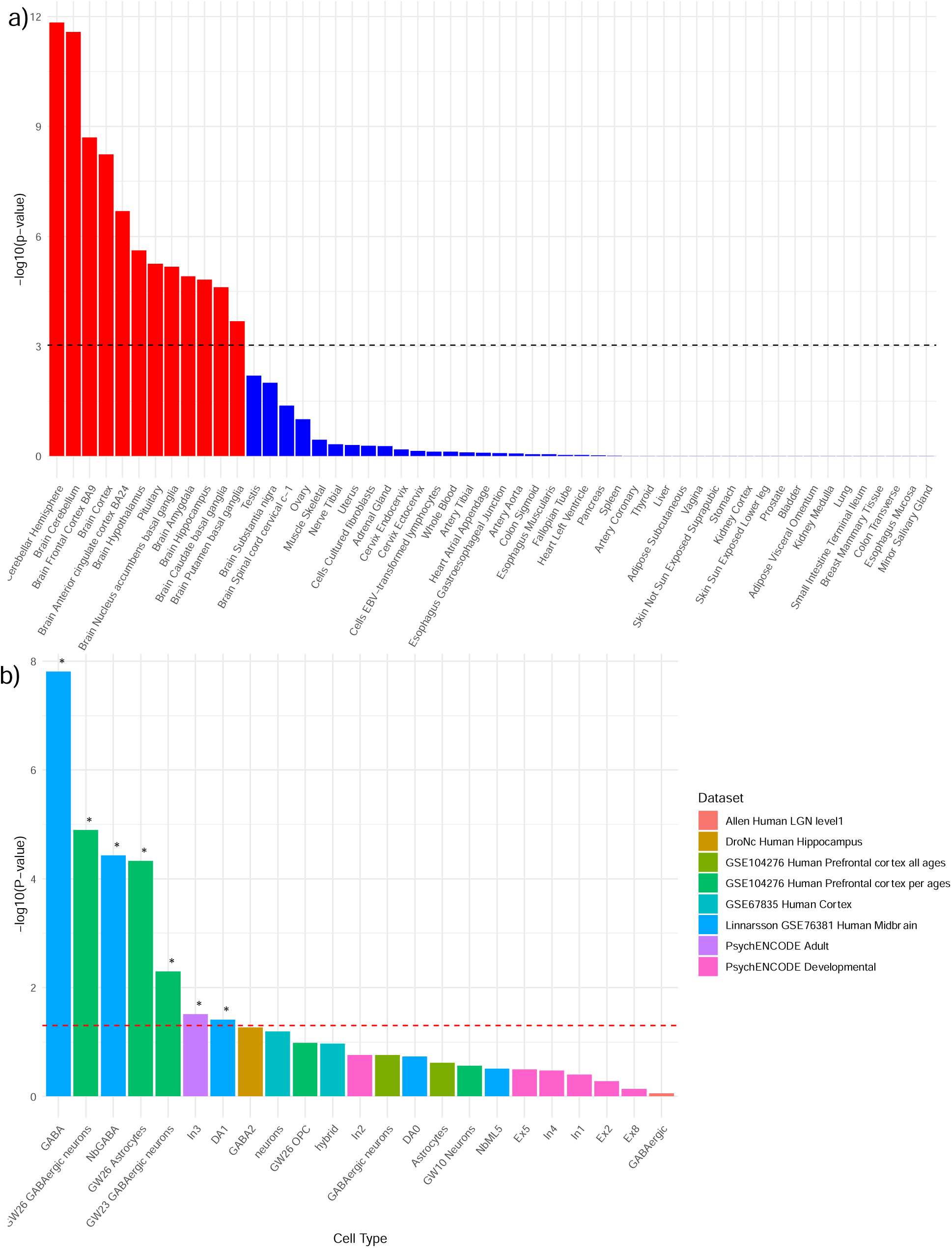
MAGMA gene-property analyses. *Note:* **a)** Gene expression in 54 GTEx tissues and **b)** cell-type specificity results.

To investigate cell-specific enrichment, we performed cell-type specificity analyses using 16 human brain scRNA-seq datasets and identified 7 cell-specific gene expression profiles in association with the somatoform factor after multiple testing correction (Figure 3b). Of the 7 cell types, 3 (GABAergic neurons in the prefrontal cortex at gestational week 26, GABAergic neurons in the human midbrain, and inhibitory neurons from PsychENCODE adult brain samples) were independently associated with the somatoform factor, with the others jointly explained by their association with the independent cell types. In cross-dataset conditional analyses, GABAergic neurons at gestational week 26 and inhibitory adult neurons were significantly collinear, suggesting that the associations of these two cell types are driven by similar genetic signals.

### SMR with Brain eQTLs and Blood Plasma pQTLs

We conducted summary-data-based Mendelian randomization (SMR) analyses to examine whether the associations of SNPs with somatoform traits were mediated by effects on gene expression in the brain and protein expression in blood plasma. We identified 28 genes whose expression levels in the brain exerted putatively causal effects on somatoform traits (Bonferroni-adjusted *p* < 0.05 and *p*_SMR_ > 0.05; Figure 4a). Among these were *UHRF1BP1*, which is known to interact with a key regulator of DNA methylation and is involved in cell apoptosis, and *HLA-DRB1*, part of the human leukocyte antigen (HLA) family of genes that is critical in initiating immune responses and implicated in many autoimmune conditions.

**Figure 4.**
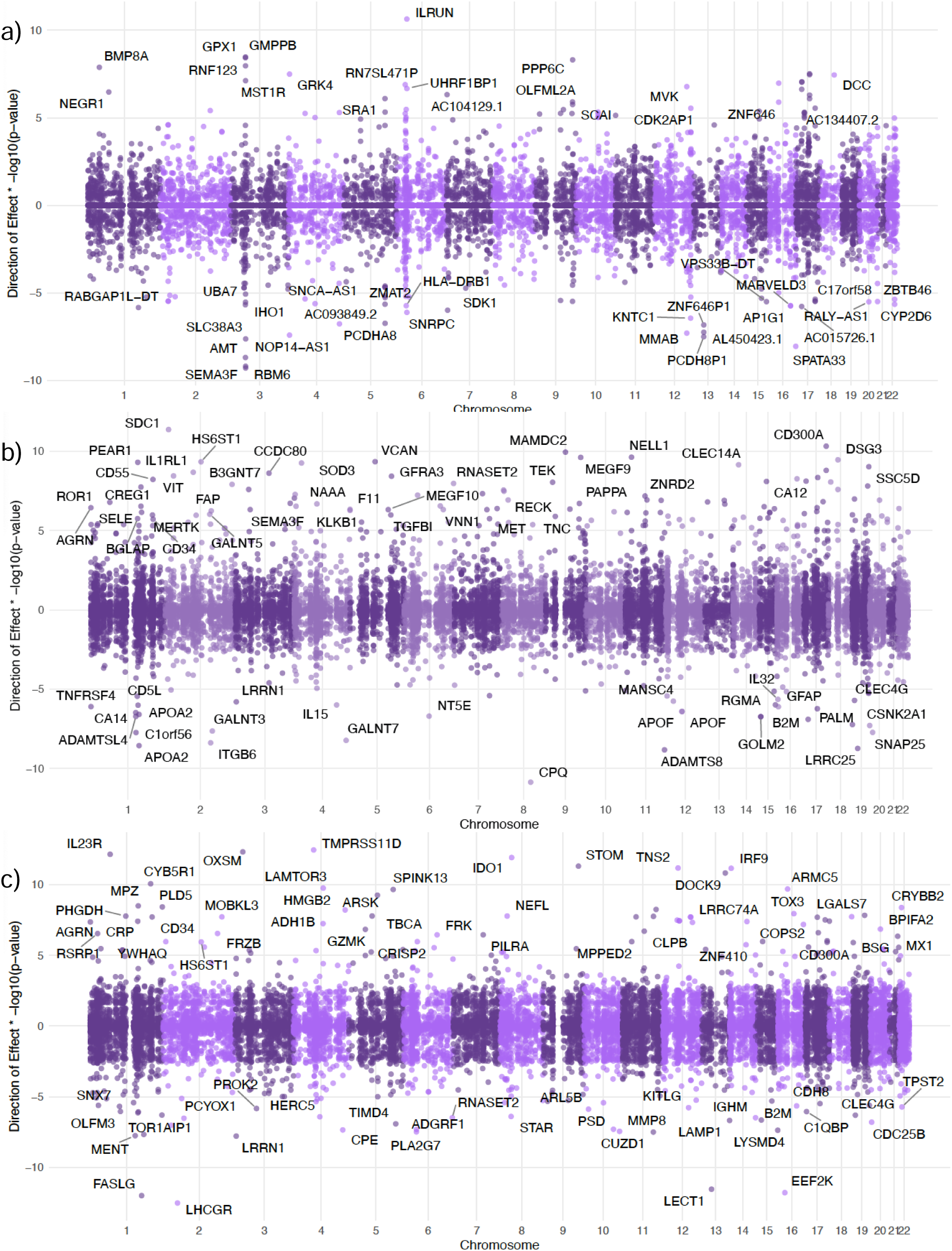
Brain eQTL and blood pQTL associations using summary-data-based Mendelian randomization. *Note:* **a)** MetaBrain eQTLs, **b)** UKB blood pQTLs, and **c**) deCODE blood pQTLs.

Using data from the UKB Pharma Proteomics Project (UKB-PPP), we identified 117 genes whose protein levels were significantly associated with somatoform traits, including several members of the CD300 family of genes that are involved in modulating inflammatory responses (Figure 4b). Additionally, genes involved in neural development and synaptic processes were significant, such as the *HS6ST1* and *LRRN1* genes. Using plasma proteomic data from deCODE, we identified 57 genes whose effects on protein abundance were putatively causally associated with somatoform traits (Figure 4c). Across the UKB-PPP and deCODE analyses, 6 genes were consistently identified: two members of the CD300 family of genes (i.e., *CD300A* and *CD300C*), *CLEC4G*, *HS6ST1*, *LRRN1*, and *RNASET2*.

### Transcriptome-Wide Association Analyses

To examine genes whose expression in enriched tissues was related to somatoform traits, we performed two TWAS using MetaXcan.^44,45^ In the first, we used S-MultiXcan to simultaneously examine gene expression profiles across the 12 tissues that showed significant enrichment in MAGMA analyses. After Bonferroni correction (*p* = 0.05/14,368 = 3.48e-6), we identified 158 genes with significantly altered expression levels associated with somatoform traits (Figure 5a). In a second TWAS of brain tissues in psychiatric cases and controls (n = 1,695),^70^ we identified 131 genes that showed significantly different (*p* ≤ 4.16e-6) expression levels (Figure 5b). Across the two TWAS, 34 genes were consistently identified, including genes related to immune function (e.g., *MST1R*), oxidative stress response (e.g., *GPX1*), neural development (e.g., *DPYSL5* and *SORCS3*), and neurotransmitter signaling (e.g., *GRK4*). Several of these genes showed enriched expression across brain tissues (e.g., *DPYSL5* and *GPX1*; Supplementary Figure 5). Across the two TWAS and the SMR analysis, five genes (*CCDC144CP*, *PPP6C*, *SCAI*, *UHRF1BP1*, *USP32P3*) were consistently implicated. **Polygenicity, Discoverability, and Polygenic Overlap with Psychopathology**

**Figure 5.**
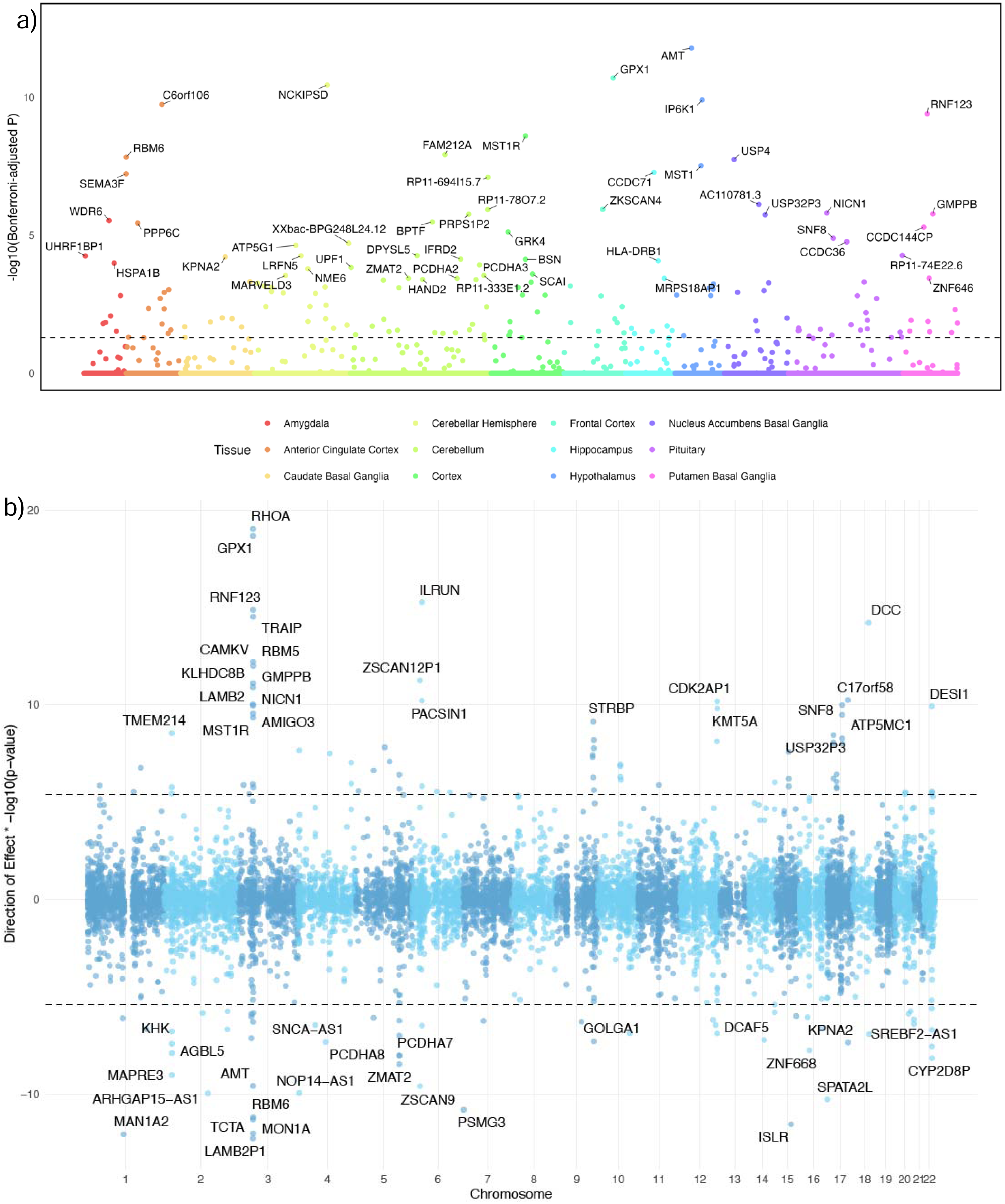
Transcriptome-wide association studies. *Note:* **a)** results from S-MultiXcan using GTEx data from 12 brain tissues, **b)** results from S-PrediXcan using PsychENCODE data from brain tissues. The top associations are annotated. Dashed lines indicate the significance threshold.

We used MiXeR software to conduct univariate and bivariate causal mixture models to assess the polygenicity (i.e., the number of variants estimated to be needed to explain 90% SNP-heritability) and discoverability (i.e., the causal effect size variance) of somatoform traits, as well as their polygenic overlap with psychopathology spectra (Supplementary Figure 6). MiXeR models estimated 11,321 causal SNPs for the somatoform factor (SD = 562.64), with an average discoverability of 7.50e-6 (SD = 3.55e-7). The somatoform factor and externalizing psychopathology were moderately genetically correlated (*r_g_* = 0.46, SD = 0.02), and shared an estimated 76% of their causal variants (SD = 0.18). Similarly, the somatoform factor and internalizing psychopathology were significantly genetically correlated (*r_g_* =0.63, SD = 0.01) and shared 79% of their causal variants (SD = 0.12). Estimates were also similar for the polygenic overlap between general psychopathology and the somatoform factor (*r_g_* = 0.69, SD = 0.01, Dice = 0.83, SD = 0.07).

### Genetic Correlations

Genetic correlations were performed between the somatoform factor and 1,426 publicly available GWAS. After Bonferroni correction (*p* = 0.05/1,426 = 3.51e-5), the somatoform factor was significantly genetically correlated with 646 phenotypes (Supplementary Table 6 and Supplementary Figure 7). Consistent with the somatoform factor representing persistent physical symptoms for which there is no identifiable medical cause, one of the top genetic correlations was with “Symptoms, signs and abnormal clinical and laboratory findings, not elsewhere classified” (r_g_ = 0.80, SE = 0.02, *p* = 1.69e-242). Of the significant associations, at least 70 (10.84%) were with pain-related conditions and medications. Many psychiatric traits were also significantly genetically correlated with the somatoform factor, including major depressive disorder (r_g_ = 0.60, SE = 0.02, *p* = 3.29e-212), mood swings (r_g_ = 0.60, SE = 0.02, *p* = 1.73e-166), and loneliness/isolation (r_g_ = 0.62, SE = 0.02, *p* = 6.02e-136). Results identified significant genetic correlations with obesity-related phenotypes (e.g., waist circumference, body mass index, and whole-body fat mass), socioeconomic status (e.g., receiving disability, educational attainment, unemployment, and financial difficulties), and general health (e.g., having a longstanding illness, taking prescription medications, lacking physical activity, and receiving a diagnosis of a serious medical condition/disability).

### Lab-and Phenome-Wide Association Scans

*BioVU and PMBB Meta-Analysis.* After meta-analyzing effects across BioVU and PMBB, there were 229 significant associations with the somatoform PGS (Figure 6). The top associations included obesity (beta = 0.18, SE = 0.01, *p* = 2.76e-76), tobacco use disorder (TUD; beta = 0.18, SE = 0.01, *p* = 9.69e-72), type 2 diabetes (beta = 0.17, SE = 0.01, *p* = 3.67e-69), and mood disorders (beta = 0.14, SE = 0.01, *p* = 3.00e-52). There were many significant associations with pain-related conditions, including nonspecific chest pain (beta = 0.12, SE = 0.01, *p* = 2.36e-38), unspecified muscle pain (beta = 0.18, SE = 0.02 *p* = 3.64e-29), abdominal pain (beta = 0.09, SE = 0.01, *p* = 2.18e-27), and chronic pain (beta = 0.12, SE = 0.01, *p* = 2.83e-22). Study-specific results from the BioVU and PMBB PheWAS are in Supplementary Tables 7 and 8.

**Figure 6.**
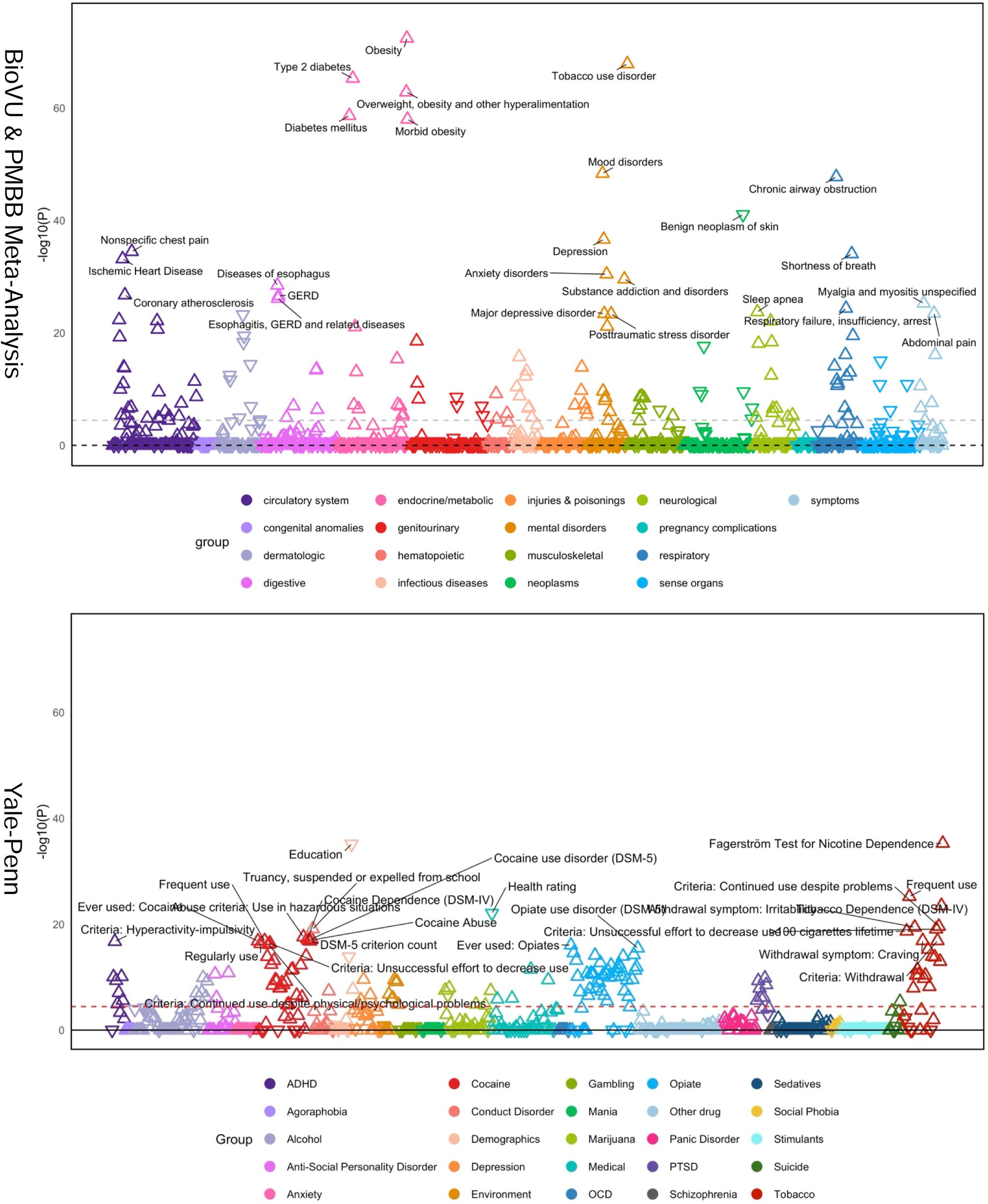
Phenome-wide association studies in BioVU, Penn Medicine BioBank, and Yale-Penn cohorts.

*BioVU LabWAS*. After Bonferroni correction, LabWAS analyses identified 40 significant associations of the somatoform factor with biomarkers (Supplementary Figure 8), including elevated C reactive protein levels (CRP; beta = 0.03, SE = 0.005, *p* = 4.43e-15) and erythrocyte sedimentation rates (beta = 0.06, SE = 0.01, *p* = 1.92e-06). Other notable associations were with higher erythrocyte distribution widths (beta = 0.06, SE = 0.004, *p* = 6.66e-52) and white blood cell counts (beta = 0.05, SE = 0.004, *p* = 1.58e-37), and with lower levels of iron (beta = -0.04, SE = 0.01, *p* = 1.75e-6) and Vitamin D (beta = -0.06, SE = 0.01, *p* = 5e-19). Consistent with the link with diabetes in the PheWAS, the somatoform PGS was also related to higher hemoglobin A1c levels (beta = 0.03, SE = 0.005, *p* = 4.43e-15).

*Yale-Penn.* In the Yale-Penn sample, there were 229 significant PheWAS associations after Bonferroni correction (Figure 6 and Supplementary Table 9). The sample is enriched for substance use disorders, and these were the most prevalent associations identified. However, other phenotypes like lower educational levels (beta = -0.17, SE = 0.01, *p* = 1.15e-38), poor self-reported health rating (beta = -0.18, SE = 0.02, *p* = 1.29e-25), lower household income (beta = -0.32, SE = 0.04, *p* = 2.19e-17), more emergency room visits (beta = 1.18, SE = 0.15, *p* = 5.13e-15), and greater childhood adversity (beta = 0.28, SE = 0.04, *p* = 1.16e-12) were also significant in the deeply phenotyped sample.

### Drug Repurposing

We used LINCS to match medication signatures to somatoform gene expression signatures. After Bonferroni correction, we identified 324 perturbagens (Supplementary Table 10) comprising a wide array of mechanisms and classes across both emerging (i.e., undergoing clinical trials; n = 113) and FDA-approved (n = 211) therapeutics. Several perturbagens were of relevance to PPS, including analgesics (e.g., diclofenac, ibuprofen, and venlafaxine), antidiarrheals (e.g., loperamide), and antidepressants (e.g., bupropion). Of note, two of the identified drugs targeted the *MAP2K1* gene and reversed the gene expression signature found in the PsychENCODE TWAS. Both drugs (pd-0325901 and selumetinib) are kinase inhibitors, with selumetinib having been approved to treat symptomatic plexiform neurofibromas in children with neurofibromatosis type 1, a genetic disorder that causes tumors to grow along nerves.^71^ Additionally, of the identified drugs, ten had gene targets that mapped to GWS SNPs (Supplementary Figure 9). These ten drugs included four dopamine receptor antagonists (i.e., nemonapride, melperone, benperidol, and carmoxirole) four kinase inhibitors (i.e., vemurafenib, dabrafenib, pf-562271, and sorafenib), an HDM2 antagonist (i.e., serdemetan), and a calcium channel blocker (i.e., nimodipine). After Bonferroni correction, the DRUGSETS analysis identified one significant association with Anatomical Therapeutic Chemical (ATC) code G04B (t = 3.89, *p* = 5.44e-5), which comprises urological drugs.

### Potentially Causal Effects of the Gut Microbiome

Of the 418 taxa in the MiBioGen and Dutch Microbiome Projects, 326 (130 from MiBioGen and 196 from the Dutch Microbiome Project) had genetic variants associated with their abundance at *p* < 1e-5 and were included in MR analyses. Of these, 23 (10 from MiBioGen and 13 from the Dutch Microbiome Project) exhibited putatively causal effects on the somatoform factor prior to multiple testing correction, but only four remained significant after Benjamini-Hochberg false discovery rate (FDR) correction (all from the Dutch Microbiome Project; Supplementary Figure 10). One of the significant results was based on a single SNP so was not interpreted. At the species level, *Adlercreutzia equolifaciens* (beta = -0.04, SE = 0.01, *p* = 5.13e-5) and *Ruminococcus bromii* (beta = -0.05, SE = 0.01, *p* = 6.65e-4) exhibited putatively causal protective effects on the somatoform factor. Similarly, the genus *Adlercreutzia* (beta = -0.04, SE = 0.01, *p* = 5.14e-5) was also significant and potentially protective. In the MiBioGen cohort, the genus *Adlercreutzia* was not putatively causally associated with the somatoform factor. However, analyses in that cohort showed a consistent direction of effect as that seen in the Dutch Microbiome Project. In the MiBioGen cohort, the putative effects of *Ruminococcus* on the somatoform factor were in a matching direction as well, and the association with the genus was significant prior to FDR correction (beta = -0.01, SE = 0.006, *p* = 0.04). Therefore, findings in MiBioGen provide relatively modest replication of our results.

Steiger directionality tests for all significant findings supported the proposed causal direction of microbiota on the somatoform factor (all *ps* < 2.05e-5). We used the Egger intercept to evaluate confounding by pleiotropy, but due insufficient instruments, we were only able to evaluate this for *Ruminococcus bromii* (Egger intercept = -0.02, SE = 0.05, *p* = 0.76). There was no evidence for heterogeneity in the effects of instruments for *Adlercreutzia equolifaciens* (*Q*(1) = 0.02, *p* = 0.88), *Ruminococcus bromii* (*Q*(2) = 3.65, *p* = 0.16), or *Adlercreutzia* (*Q*(1) = 0.03, *p* = 0.86).

## Discussion

Although often considered distinct in clinical practice, our findings support a common latent genetic factor that contributes to vulnerability to multiple PPS, including fatigue, IBS, and pain intensity, as well as symptom perceptions, including health satisfaction and pain intensity. This aligns with the emerging view of PPS as part of a broader spectrum linked by shared etiological pathways.^72^ Amid calls for improved classification and treatment of PPS and growing recognition of the complex interactions between mind and body,^72,73^ our findings provide unique insights into the shared genetic architecture underlying these traits, opening new avenues for improved clinical care.

By incorporating various somatoform traits in an effective sample size of 799,429 EUR individuals, we identified 134 loci associated with the common genetic liability to somatoform traits, including 8 lead SNPs in novel loci not previously GWS in relation to any individual PPS. The power of the multivariate approach also allowed us to replicate findings for 44 loci that were not GWS in the input GWAS but had previously been associated with PPS. Functional annotation indicated enrichment of candidate SNPs in several regulatory regions, with the majority (59.17%) having functions related to binding and gene expression. This suggests a significant role for gene regulatory mechanisms in the etiology of PPS.

The expression of genes related to somatoform traits was enriched across 12 brain tissues, particularly the cerebellar hemisphere and cerebellum. Recently, the cerebellum has drawn attention for its potential role in modulating the emotional and cognitive elements of pain through its communication with subcortical and cortical regions.^74^ Individuals with chronic pain show altered activation patterns in the cerebellum during pain,^75^ and differences in functional connectivity between the cerebellum and other brain regions are correlated with ratings of pain intensity.^76^ Furthermore, the cerebellum may be associated with salience processing,^74,77^ which involves the integration of internal and external sensory information and has been implicated in both chronic pain^77–79^ and IBS.^80^ Cell-type specific analyses implicated the role of inhibitory GABAergic neurons in the human midbrain and prefrontal cortex. Mouse models have underscored the crucial role of GABAergic interneurons in regulating sensory sensitivity,^81^ and a GWAS of pain intensity also implicated GABAergic neurons.^9,81^ Dysregulation of inhibitory control via GABAergic neurons during development may similarly contribute to heightened sensitivity to PPS.

Performing TWAS and SMR analyses, we sought to identify potential causal genes in enriched tissues using brain transcriptomic data from PsychENCODE, GTEx, and MetaBrain. Across the two TWAS and the SMR analysis, five genes (*CCDC144CP*, *PPP6C*, *SCAI*, *UHRF1BP1*, *USP32P3*) were consistently implicated, providing strong evidence for their role in somatoform traits. Two of the identified genes (*CCDC144CP* and *USP32P3*) are pseudogenes, which have traditionally been considered non-functional but have recently been found to have important functional and regulatory roles that warrant further investigation.^82^ The remaining three genes are involved in cell signaling (*PPP6C*), cell migration and tumor suppression (*SCAI*), and regulation of DNA methylation (*UHRF1BP1*). We also identified six genes whose effects on protein levels mediated their association with somatoform traits, including four involved in immune regulation (*CD300A*, *CD300C*, *CLEC4G*, and *RNASET2*) and two in neuronal development (*HS6ST1* and *LRRN1*). Preclinical research is needed to examine the mechanisms by which these prioritized genes influence PPS.

We found substantial polygenic associations between the somatoform and internalizing, externalizing, and general psychopathology factors. Genetic correlations with 1,426 publicly available GWAS further highlighted the shared etiology of somatoform traits and psychopathology, with the genetic correlation with depression being among the strongest. This shared genetic architecture may help to explain the high comorbidity between PPS and psychopathology, particularly depression and anxiety, although our analyses do not provide information on causality or the direction of effect.^3^ In three biobanks, we found that somatoform PGS were associated with the presence of numerous physical and mental health conditions, including diabetes, TUD, mood disorders, obesity, post-traumatic stress disorder, and sleep disorders. Collectively, these findings underscore the need for treatment approaches that recognize the interconnectedness of physical and mental health.

Addressing the shared risk factors for various PPS could potentially improve outcomes across health domains. Thus, we examined potential causal effects of gut microbiota on the somatoform factor and performed drug repurposing to identify treatments relevant to a broad range of PPS. Across two independent datasets of host-gut microbiome associations, *Ruminococcus bromii* had the strongest support for potential protective effects on somatoform traits. *R. bromii* are a keystone bacterial species for their ability to metabolize resistant starch.^83^ They also contribute to butyrate production in the colon, which is used by beneficial gut microbes.^83^ A reduced abundance of *R. bromii* has been associated with chronic pancreatitis^84^ and Crohn’s disease.^85^ Importantly, even short-term dietary changes substantially alter *R. bromii* abundance in the gut,^86^ suggesting that it is a readily modifiable target. We also identified drugs that may have promise for treating multiple PPS, including 10 compounds targeting genes mapped by GWS variants. Notably, four of these compounds were dopamine receptor antagonists, including atypical antipsychotics (nemonapride, melperone, and benperidol) and a peripherally active D2 receptor antagonist with antihypertensive properties (carmoxirole).^87^ Two compounds (PD-0325901 and selumetinib) were found to reverse the brain transcriptomic expression signature of the somatoform factor by targeting the *MAP2K1* gene. One of these, PD-0325901, has potent anti-inflammatory activity.^88^ Whereas mitogen-activated protein kinase (MAPK) signaling pathways are involved in pain sensitization and their inhibition reduces pain in animal models,^89^ our findings suggest that there may be broader applications of these inhibitors in treating other PPS as well.

## Limitations

This study has several limitations. First, the summary statistics included were exclusively from EUR individuals, potentially limiting the generalizability to individuals genetically similar to non-EUR populations. Future research should aim to replicate findings in other groups as biobanks continue to grow and become more diverse. For example, a study applying gSEM in AFR individuals has shown that when GWAS are available and appropriate steps are taken to model the complex LD patterns, these models yield accurate and meaningful results.^90^ Additionally, although genetic correlation and PheWAS analyses underscored the complexity of interactions between physical symptoms and mental health conditions, they do not provide information on the mechanisms underlying these associations, which could include shared environmental risk factors, indirect or direct genetic effects, or other possibilities. Third, the associations between gut microbiota and somatoform traits, while promising, require replication and experimental research to uncover their potential mechanisms. Finally, further research is necessary to validate the efficacy and safety of the therapeutic targets identified through drug repurposing analyses, emphasizing the ongoing challenge of translating genetic discoveries into effective treatments.

## Conclusions

By identifying a common genetic factor that contributes to vulnerability to multiple PPS, our findings highlight shared biological pathways that link conditions often considered distinct in clinical practice. Enrichment analyses point to the cerebellum and GABAergic neurons as key players in the neurobiology of various PPS. The significant genetic correlations and high polygenic overlap with psychopathology align with dimensional taxonomic approaches, such as HiTOP. These findings enhance our understanding of PPS, open new avenues for research and therapeutic development, and underscore the need for approaches that address the complex interplay between genetic, neurological, and psychological factors in health and disease.

## Data Availability

GWAS Summary Statistics used for the present study can be accessed at the following locations: UK Biobank (http://www.nealelab.is/uk-biobank/), dbGaP accession phs001672 for Million Veteran Program (https://www.ncbi.nlm.nih.gov/projects/gap/cgi-bin/study.cgi?study_id=phs001672), European Bioinformatics Institute GWAS Catalog (https://www.ebi.ac.uk/gwas/) under accession number GCST90016564. Once accepted for publication, summary statistics for the somatoform factor will be made publicly available.

## Acknowledgments

Veterans Integrated Service Network 4, Mental Illness Research, Education and Clinical Center, National Institute on Alcohol Abuse and Alcoholism grants R01 AA030056 and K01 AA028292 (to RLK), and National Human Genome Research Institute grant T32 HG009495 (to KLF).

