## Supplementary Materials for "Multivariate, Multi-omic Analysis in 799,429 Individuals Identifies 134 Loci Associated with Somatoform Traits"

### Supplementary Figures

#### **Supplementary Figure 1. Genetic correlations between input traits.**

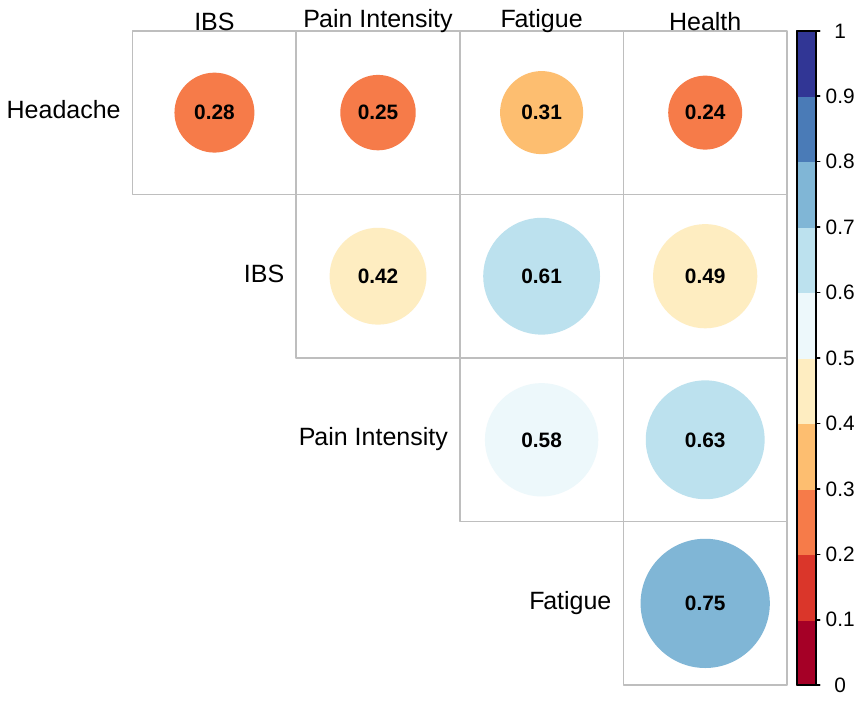

All correlations presented are significant at *p* < 0.05. IBS = irritable bowel syndrome, health = health satisfaction, reverse coded.

#### **Supplementary Figure 2. SNP-level PheWAS for lead SNPs from novel somatoform loci.**

rs10034374

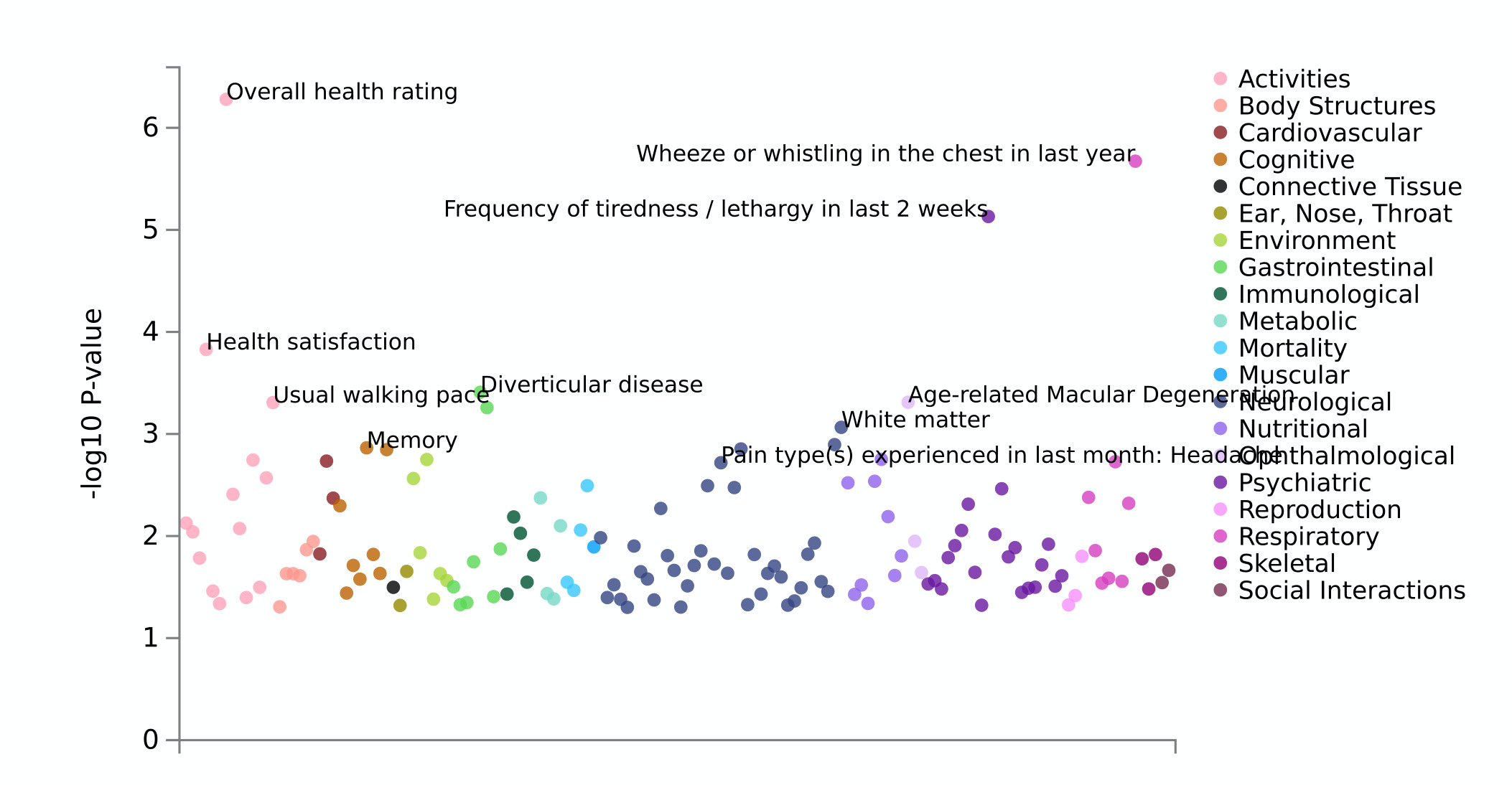

rs1514508

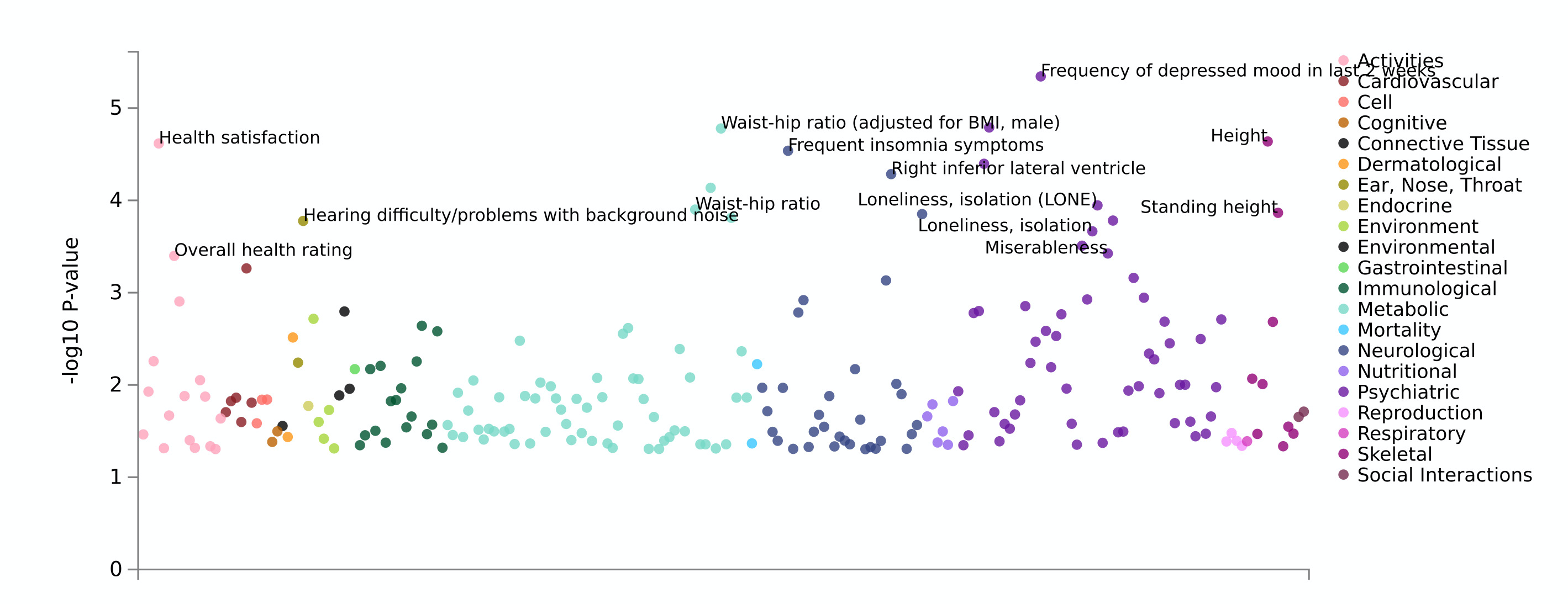

rs3003575

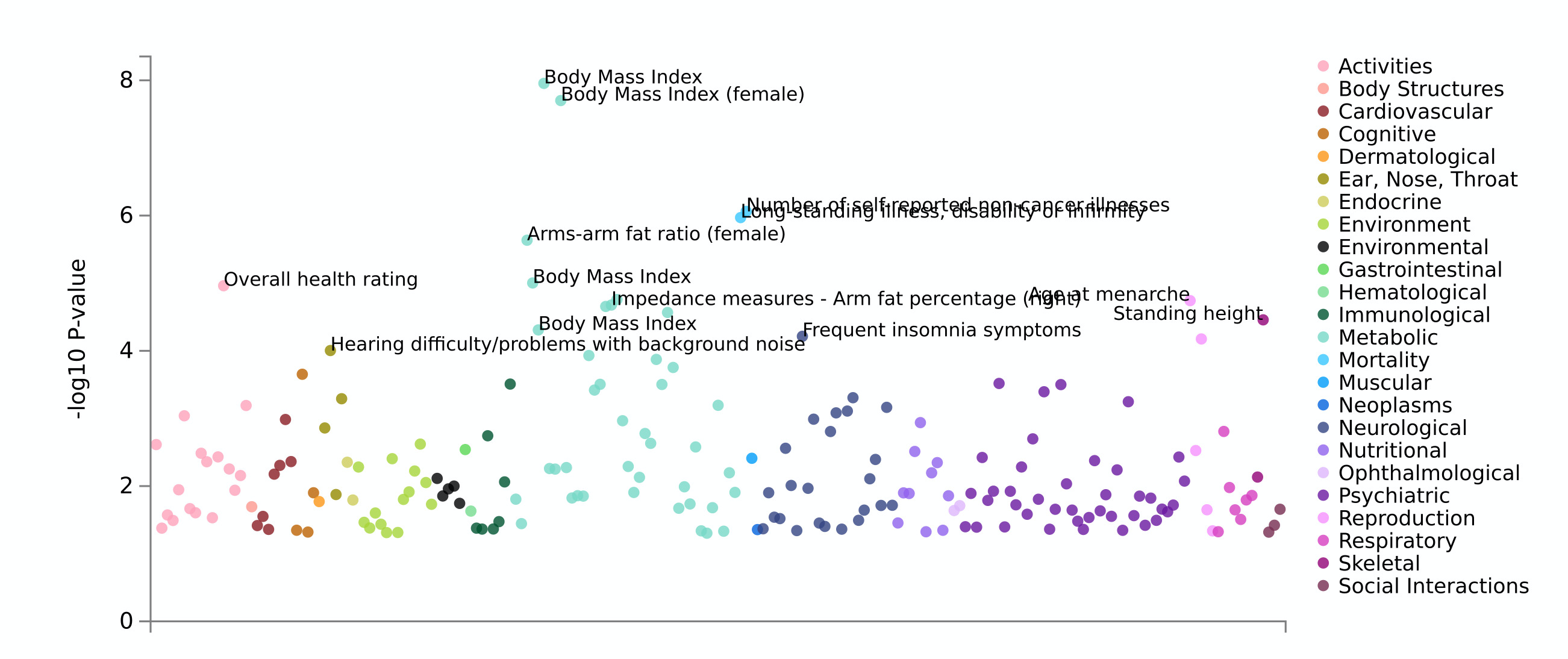

rs12232639

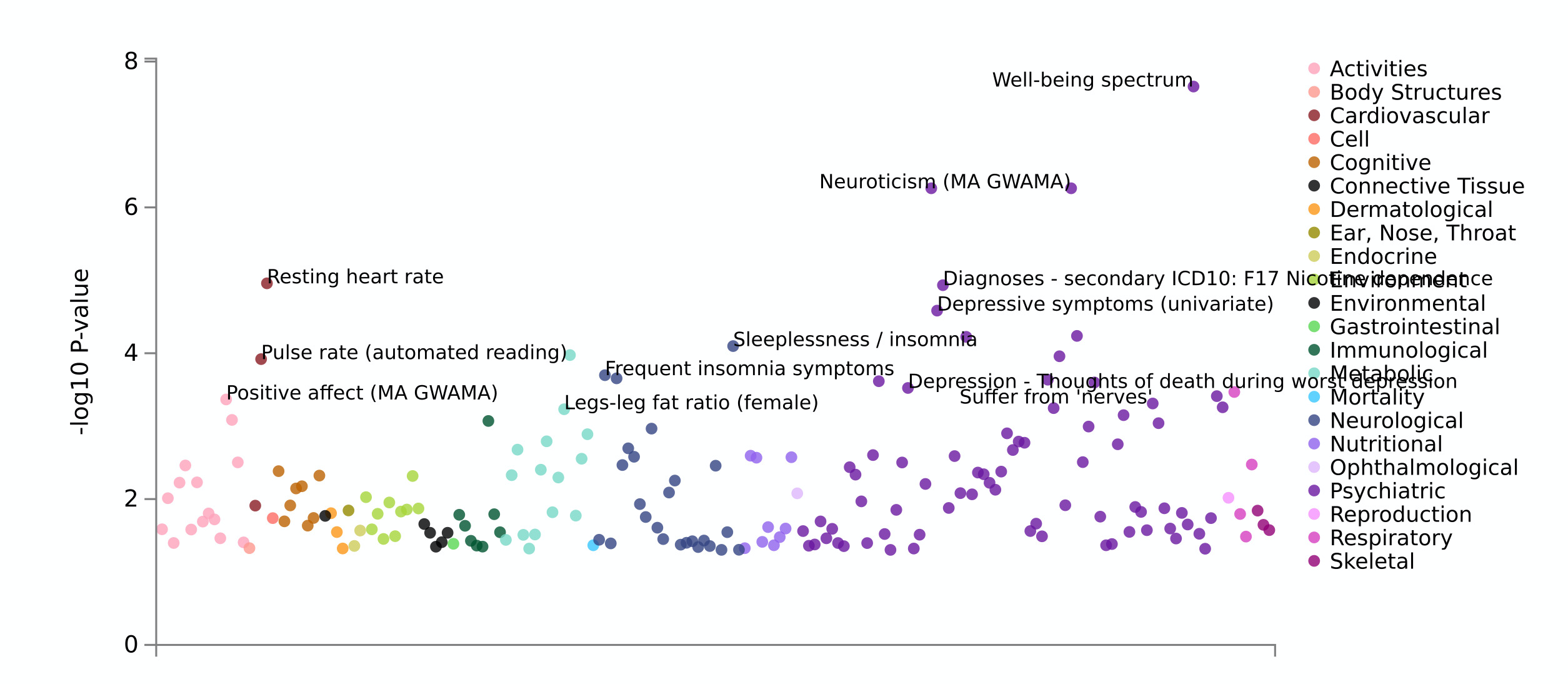

rs2879086

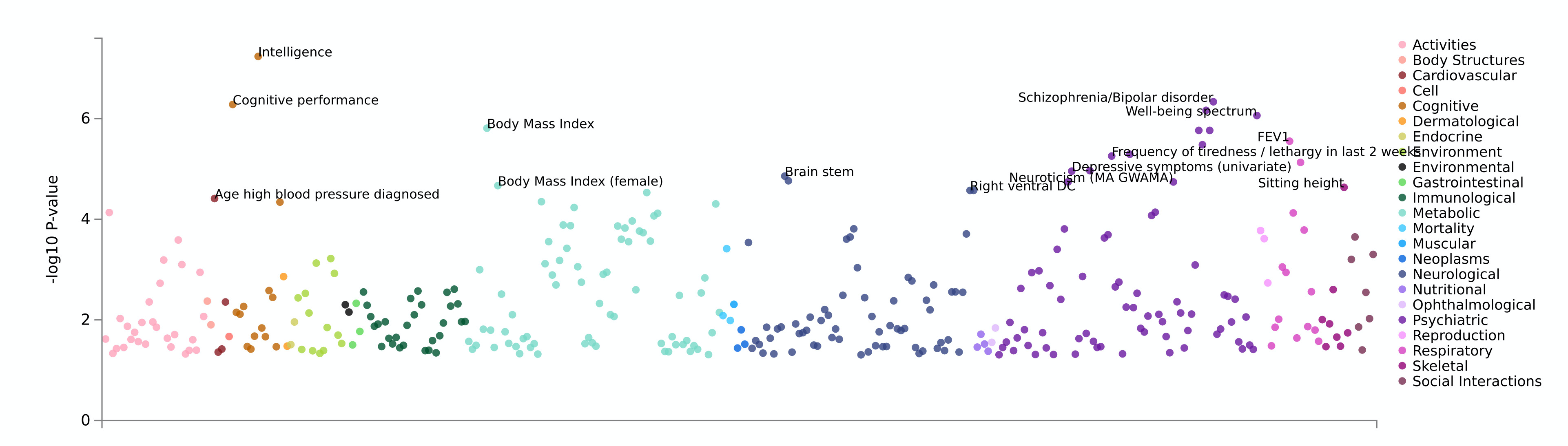

rs1351394

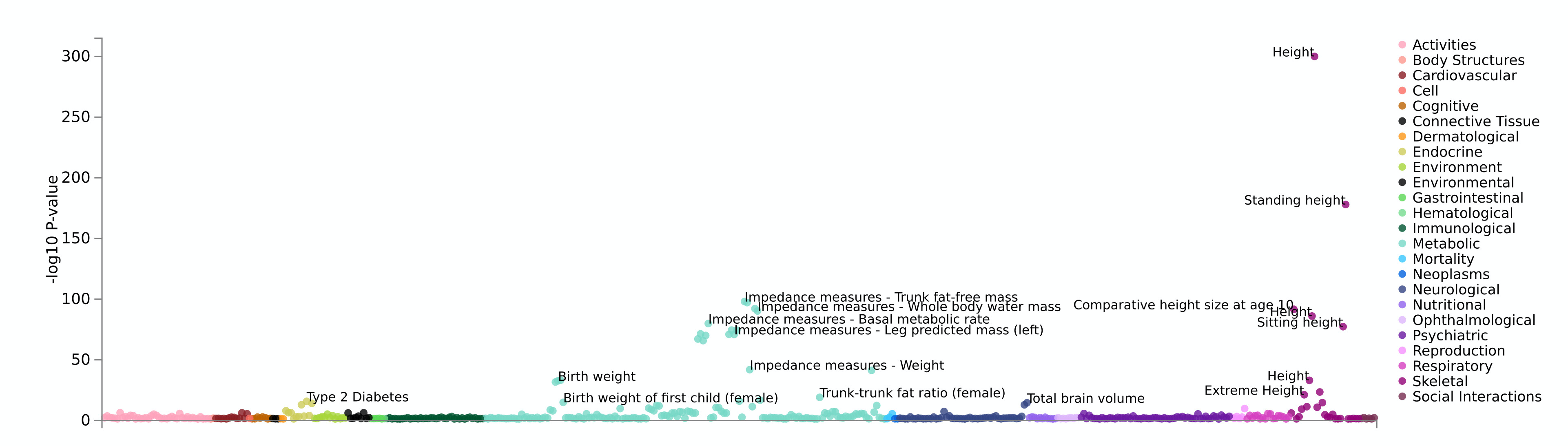

rs4696651

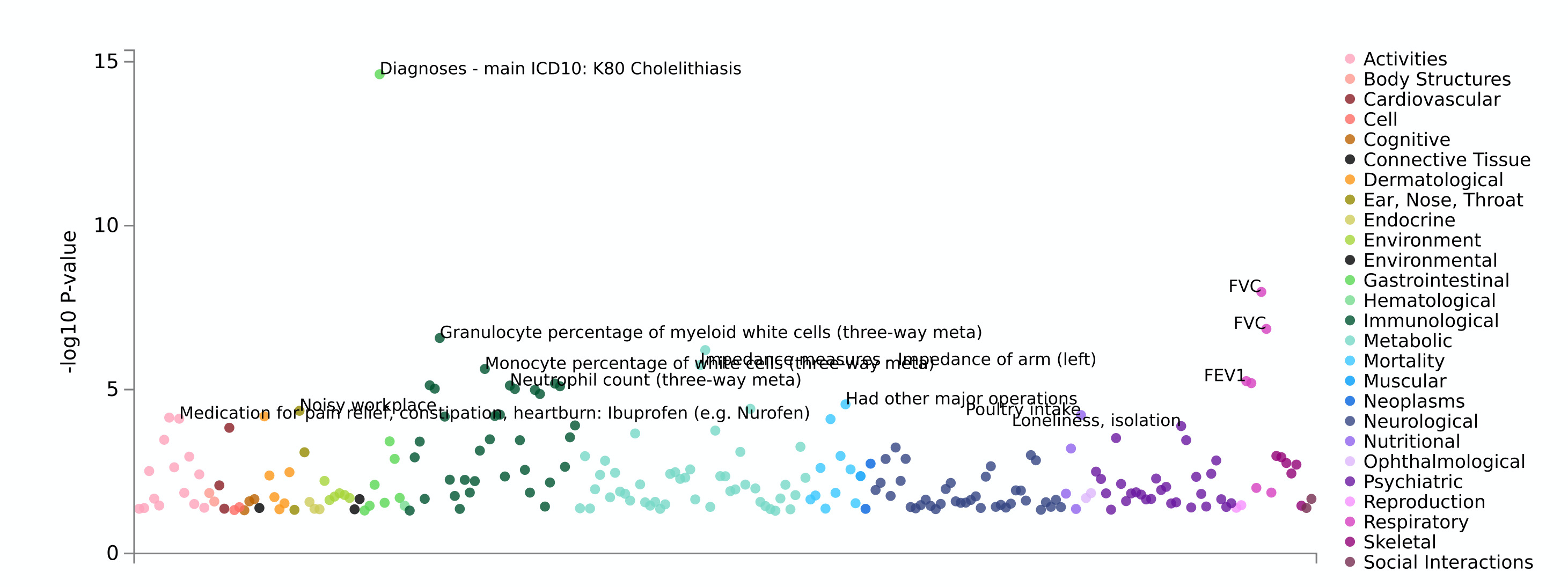

rs7640162

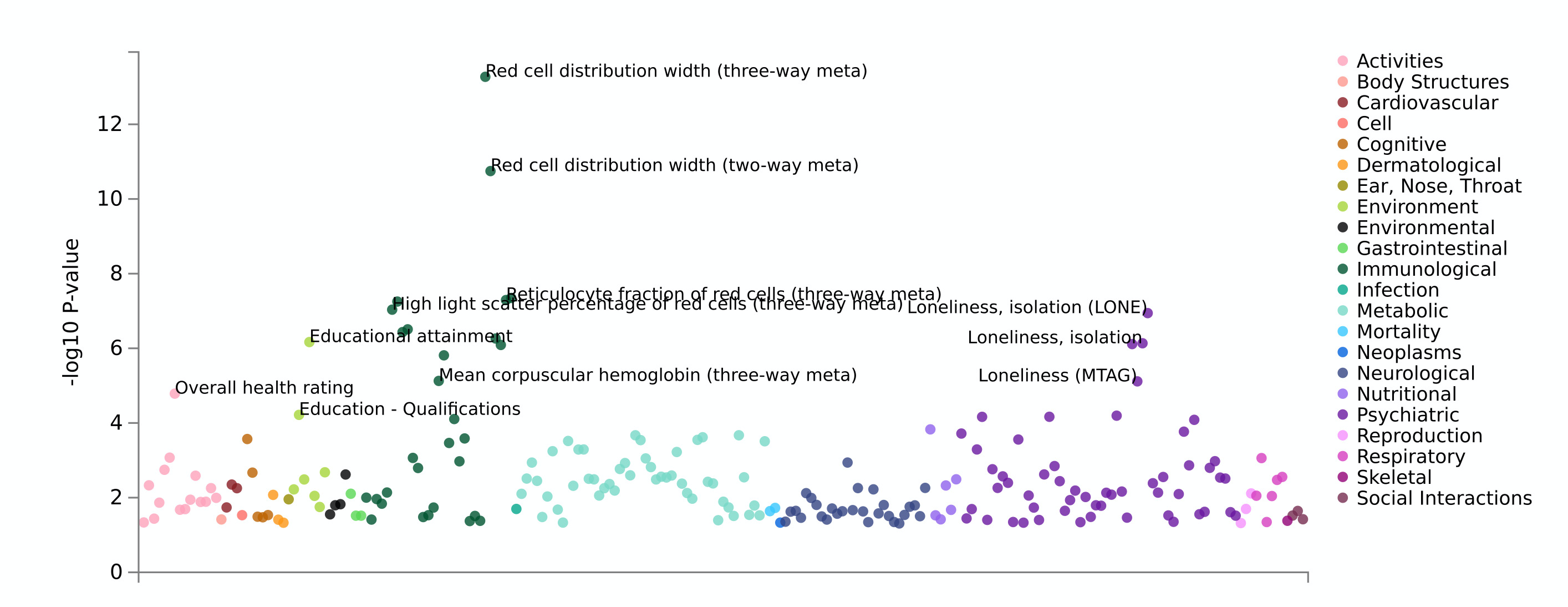

SNP-level PheWAS were performed using GWAS Atlas: <https://atlas.ctglab.nl/PheWAS>.

#### **Supplementary Figure 3. eQTL and chromatin interaction mapping of lead SNPs.**

Chromosome 1

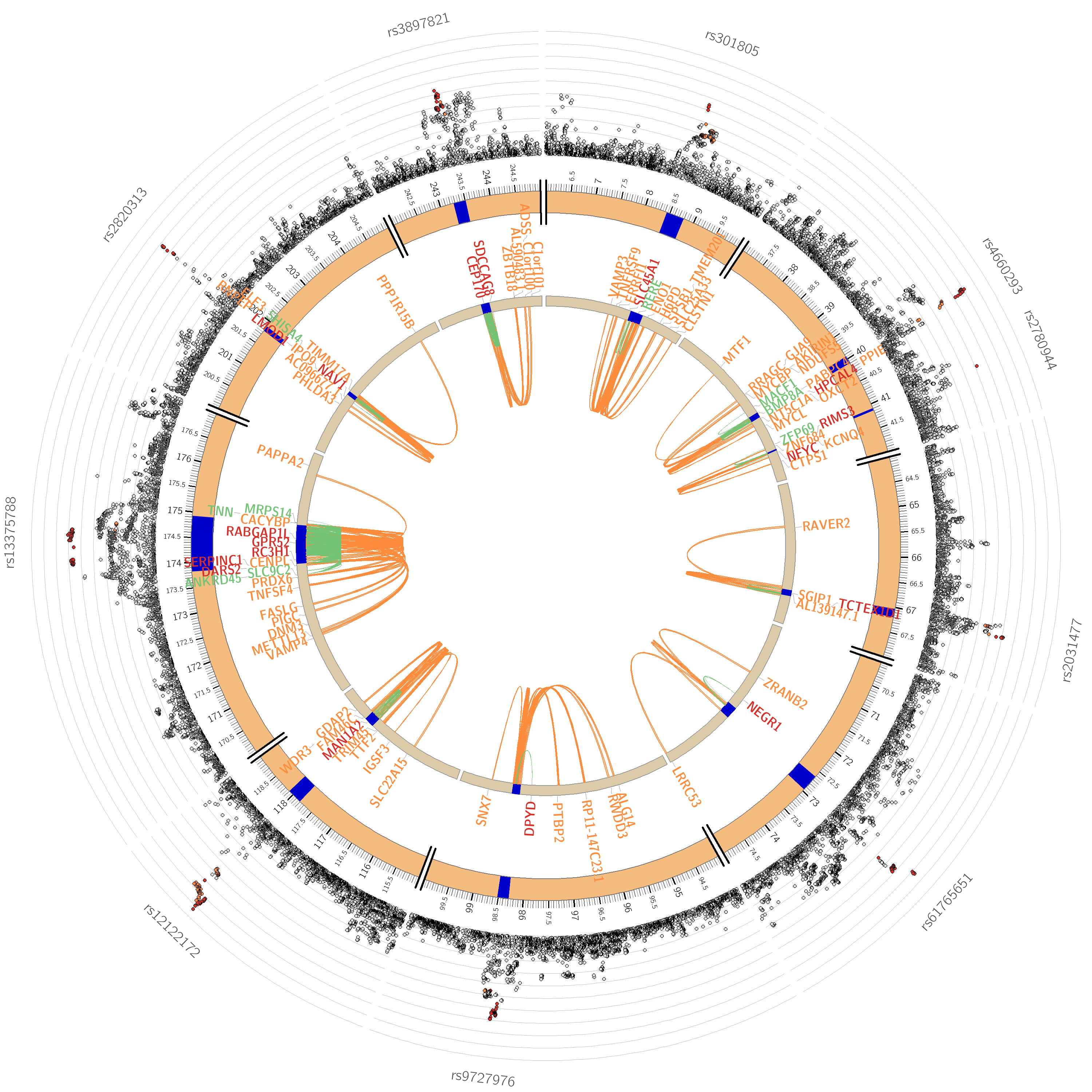

Chromosome 2

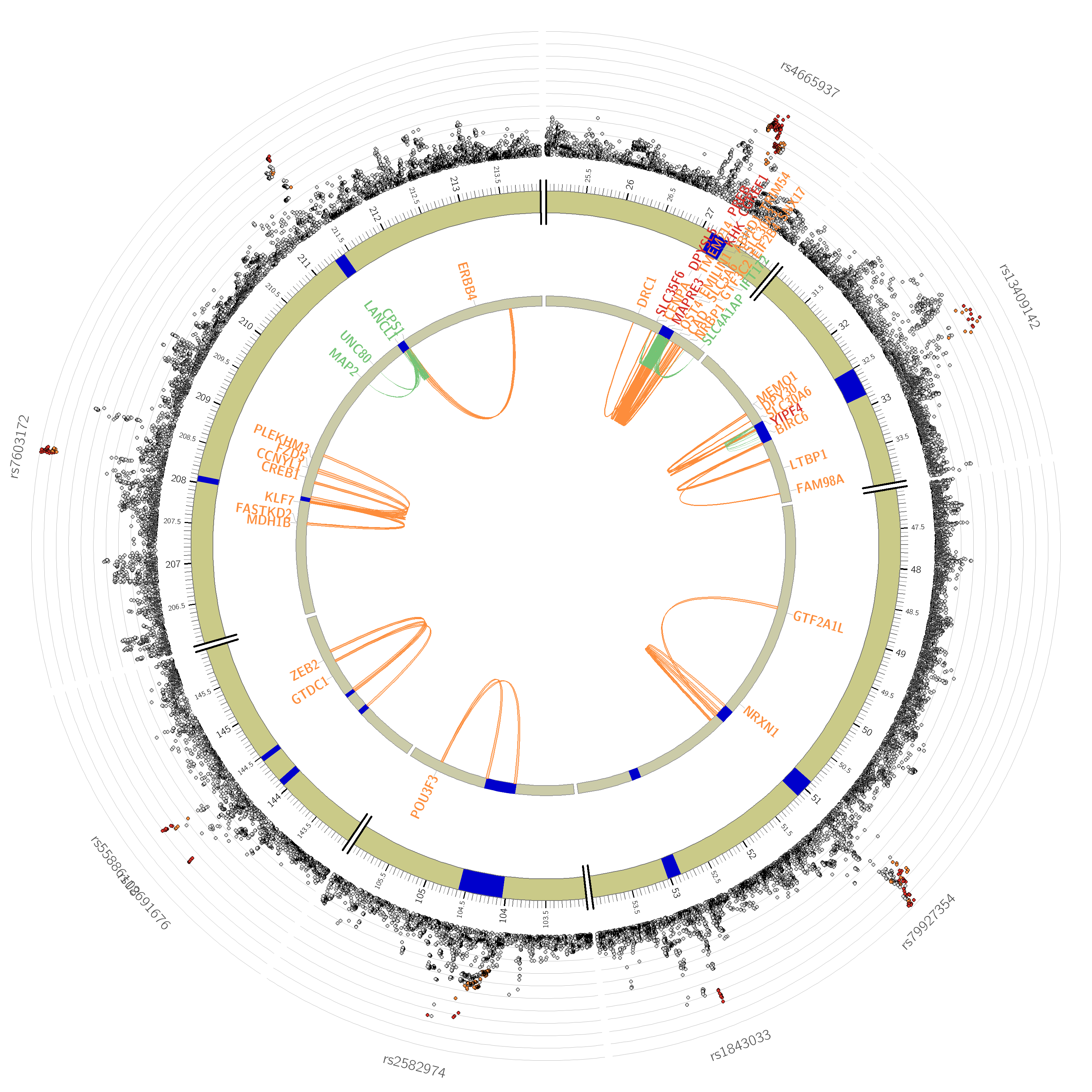

Chromosome 3

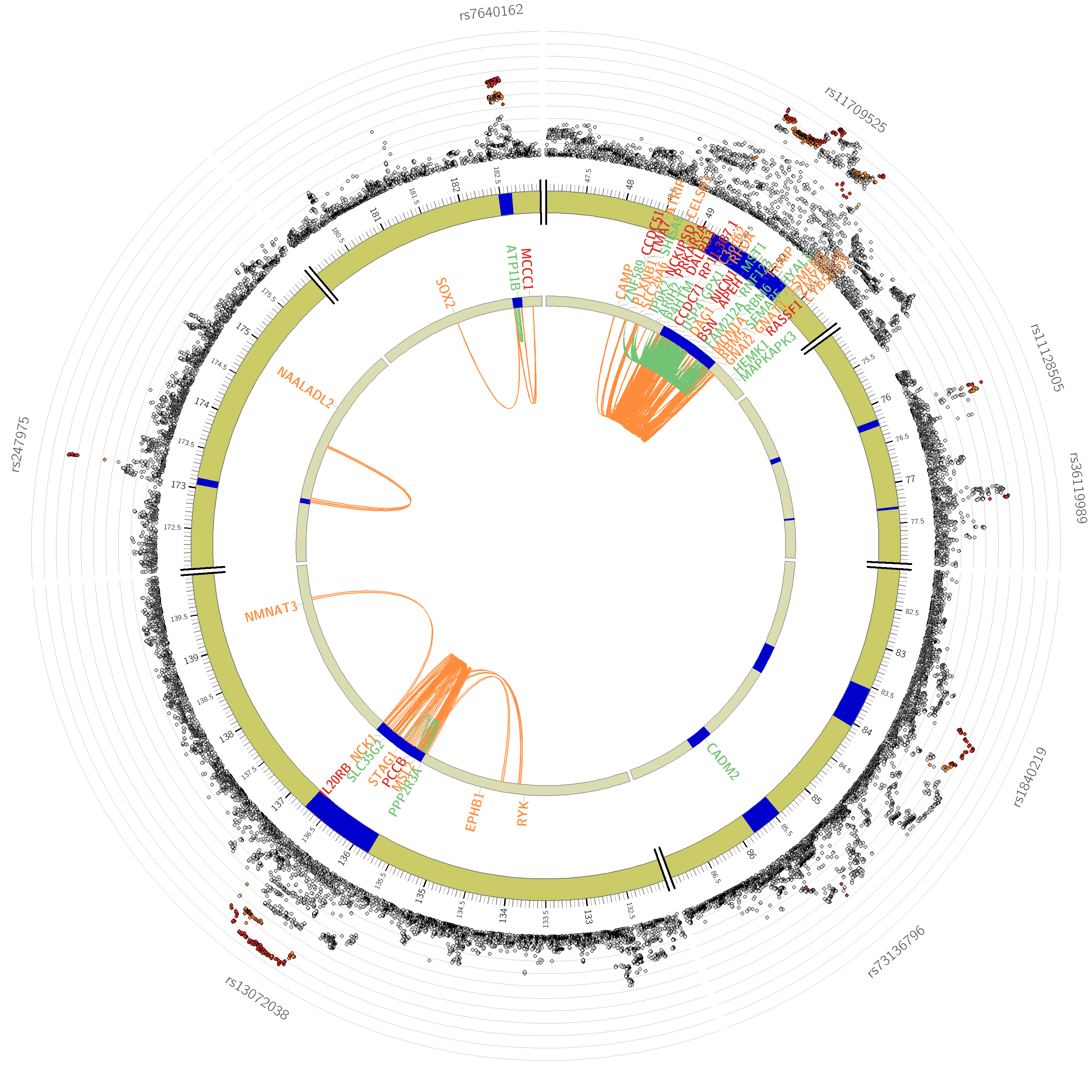

Chromosome 4

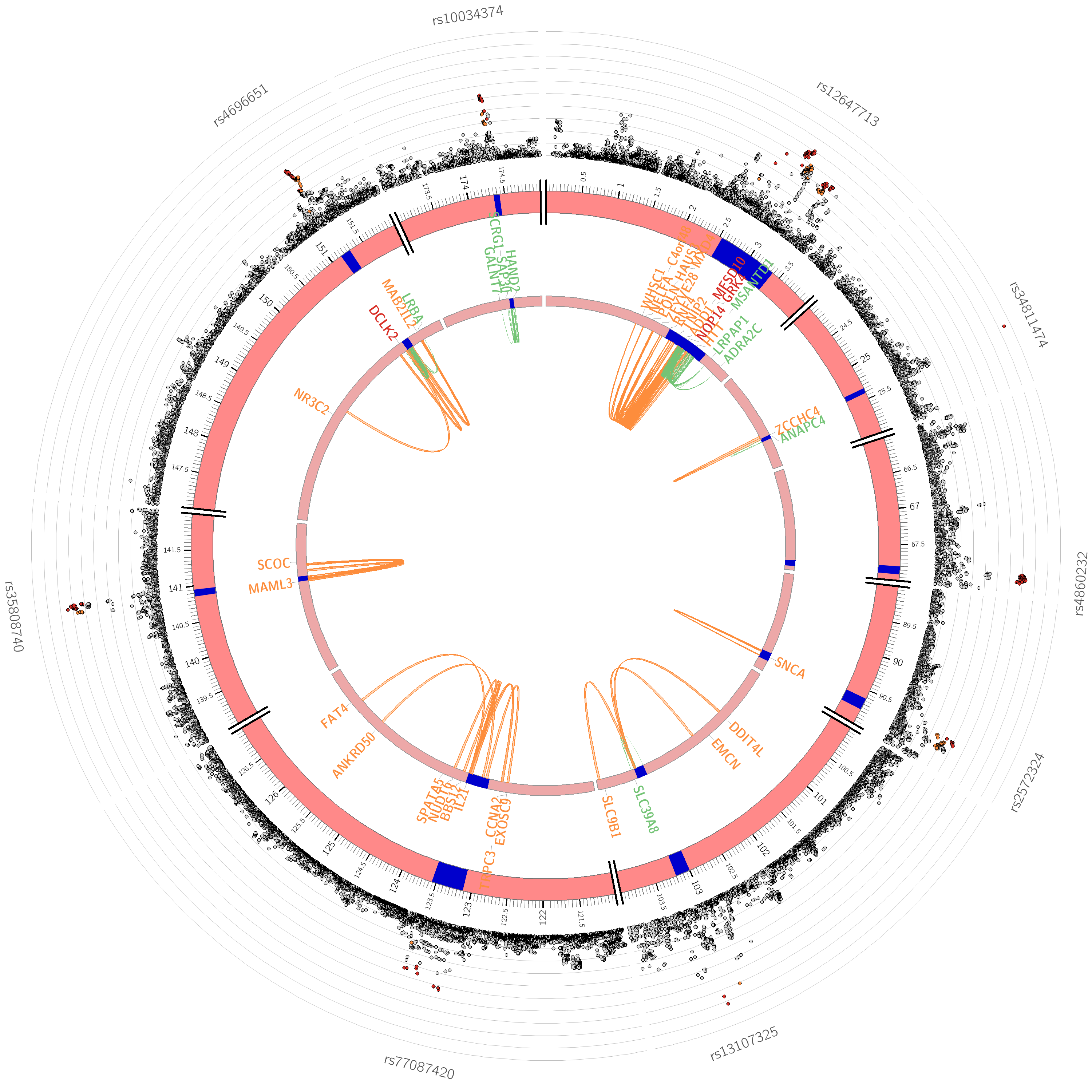

Chromosome 5

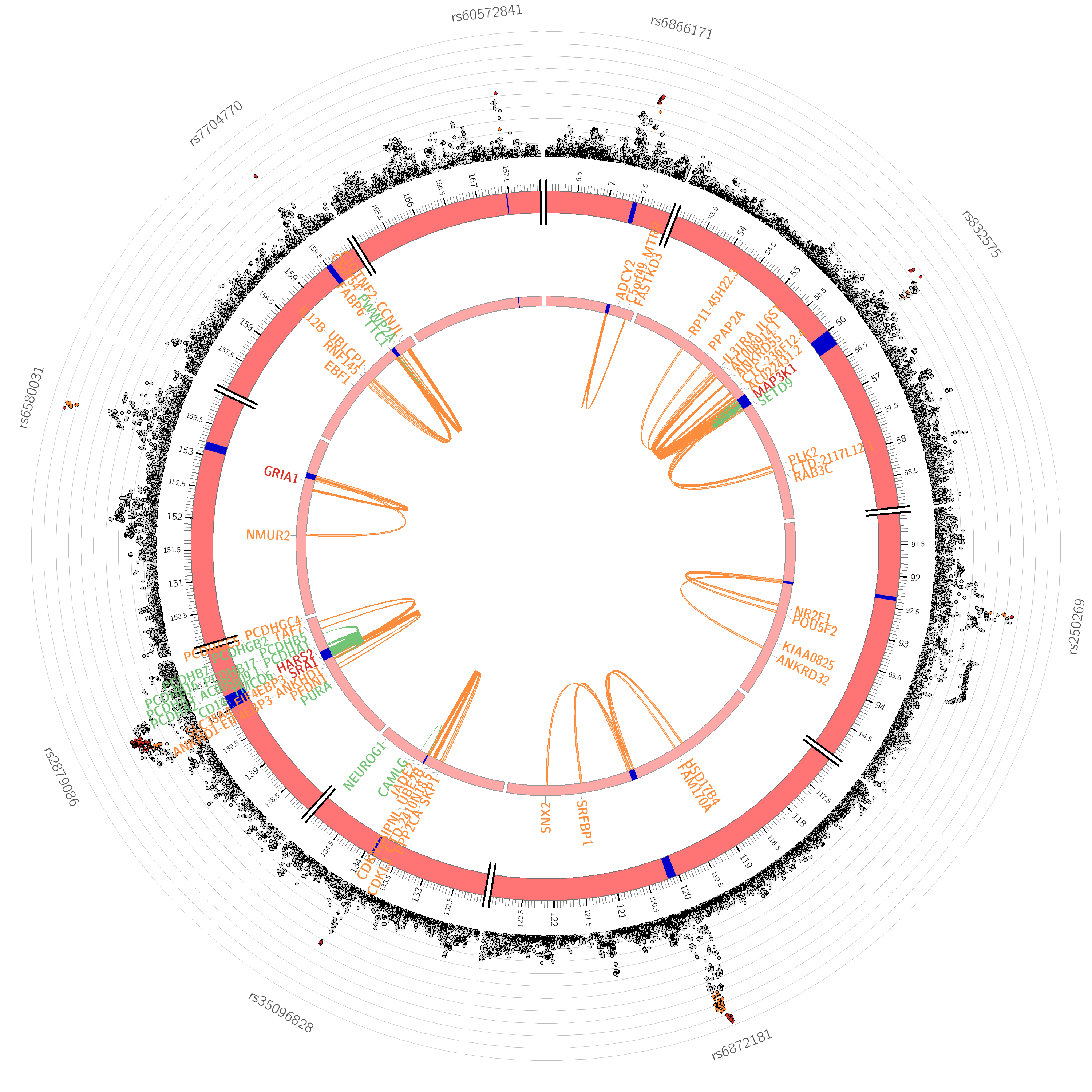

Chromosome 6

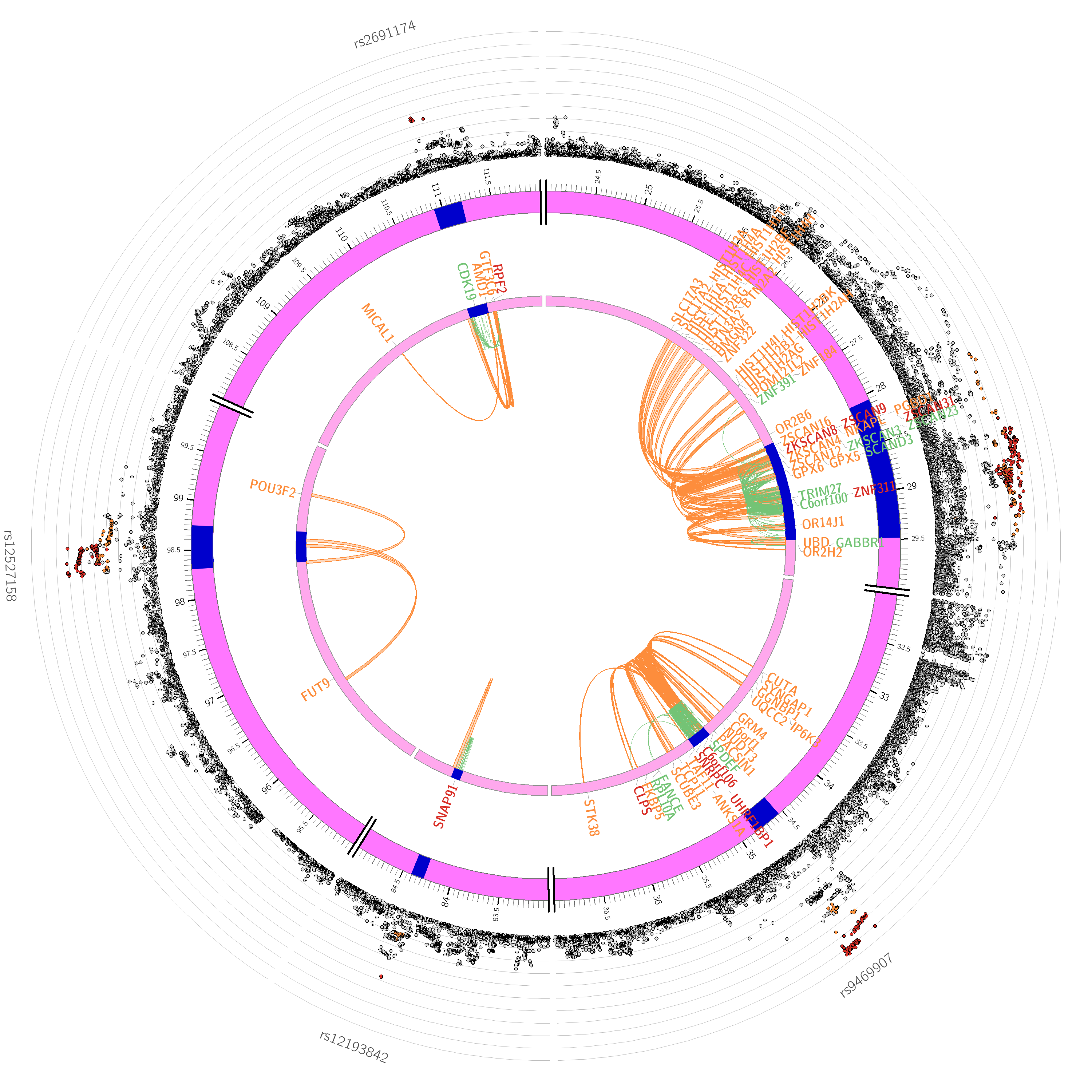

Chromosome 7

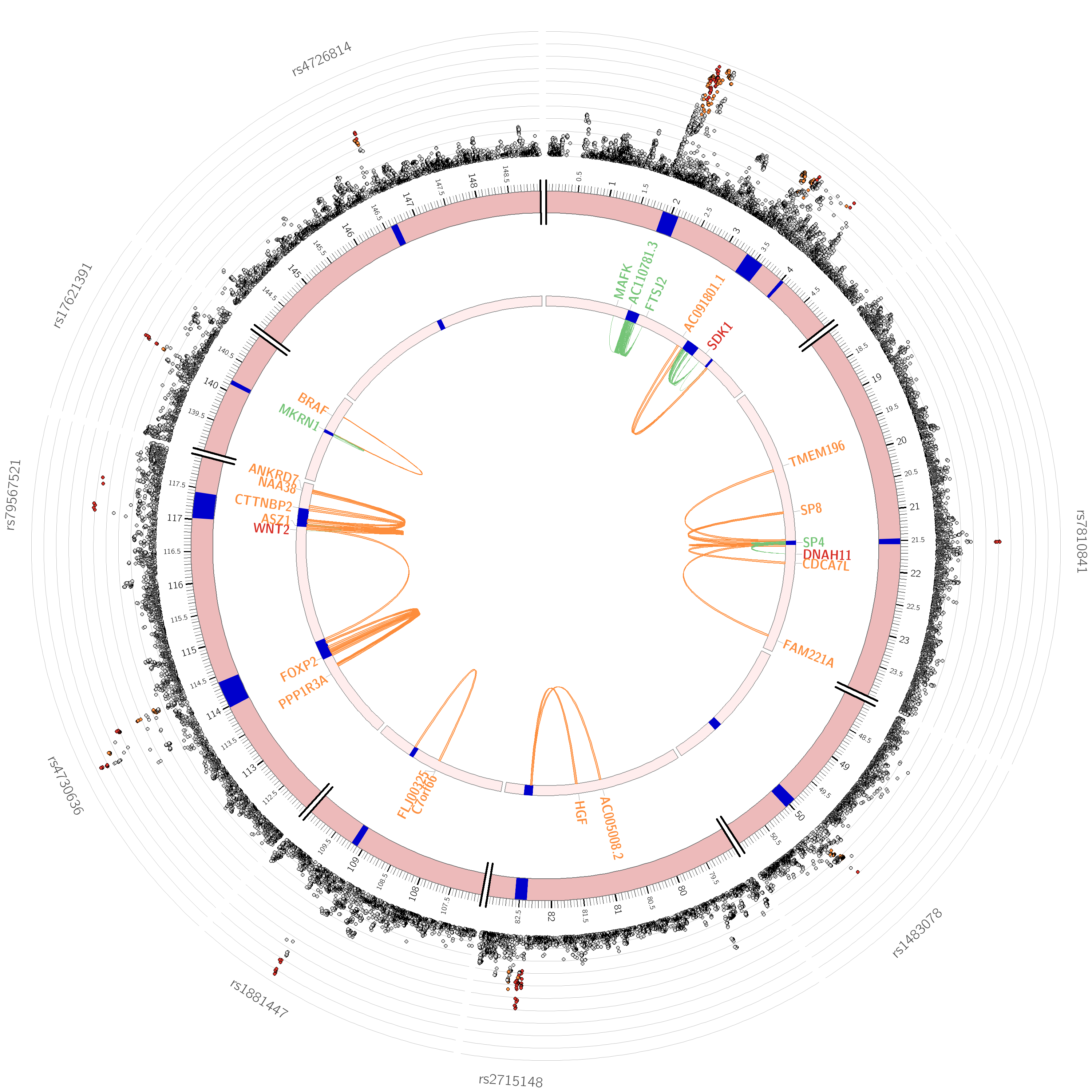

Chromosome 8

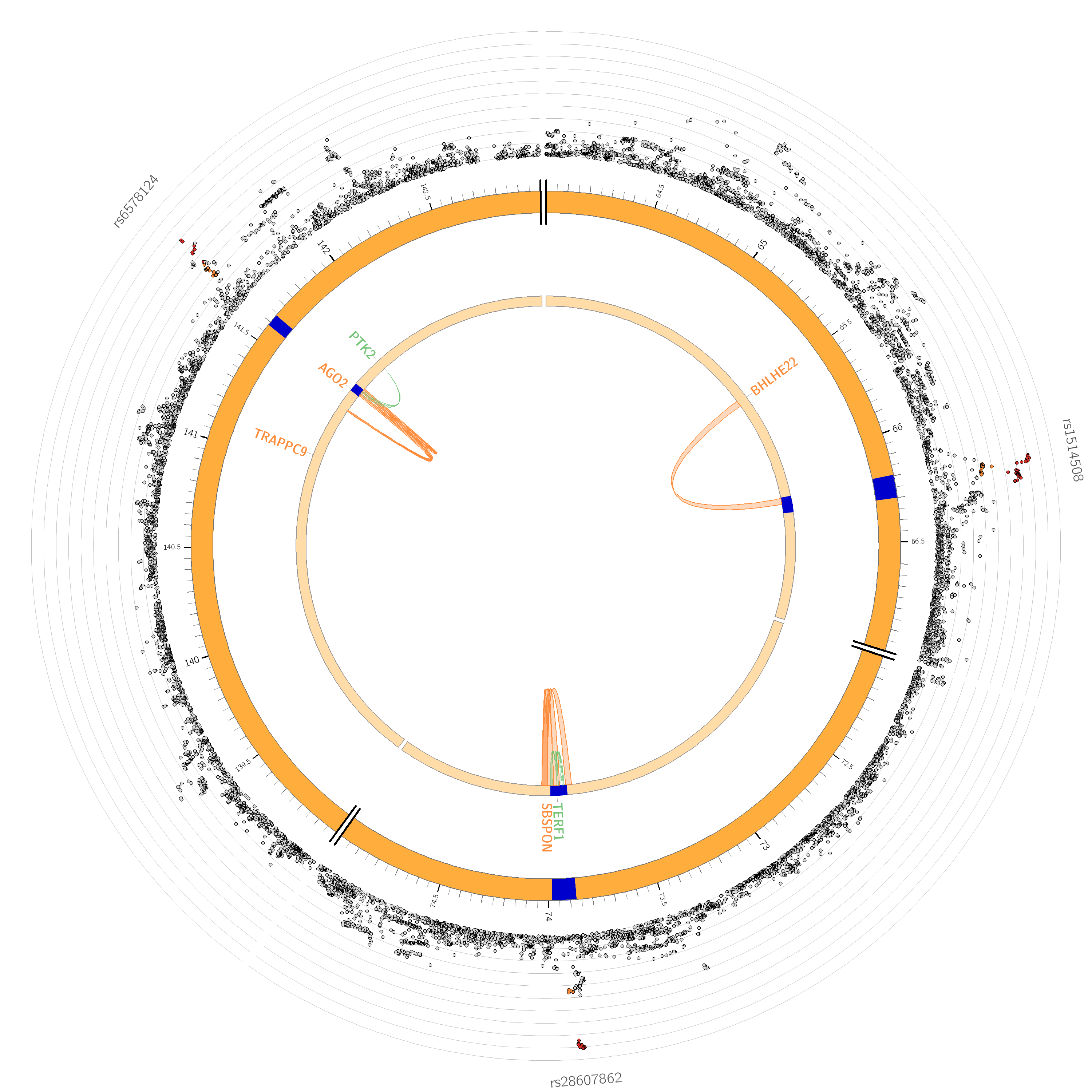

Chromosome 9

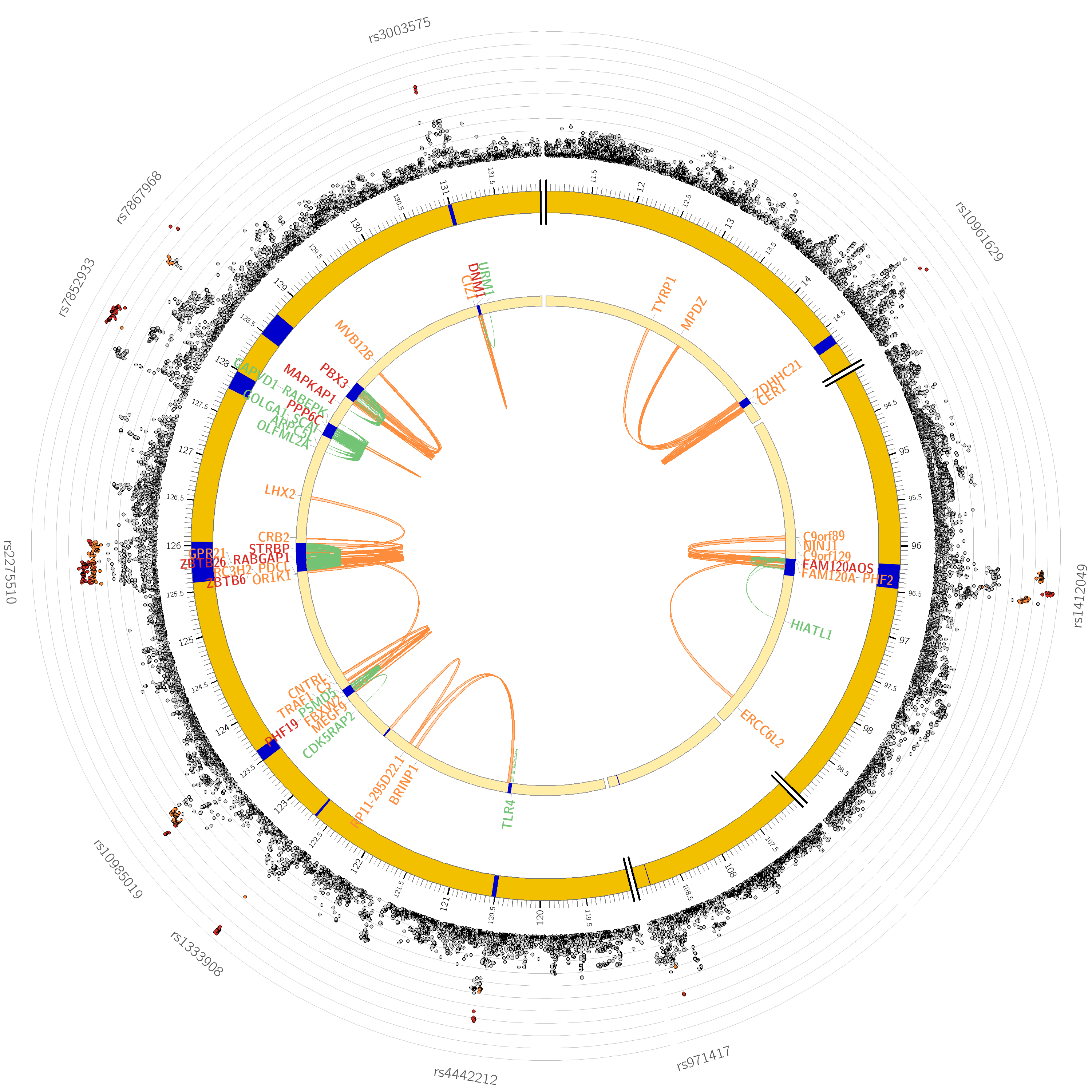

Chromosome 10

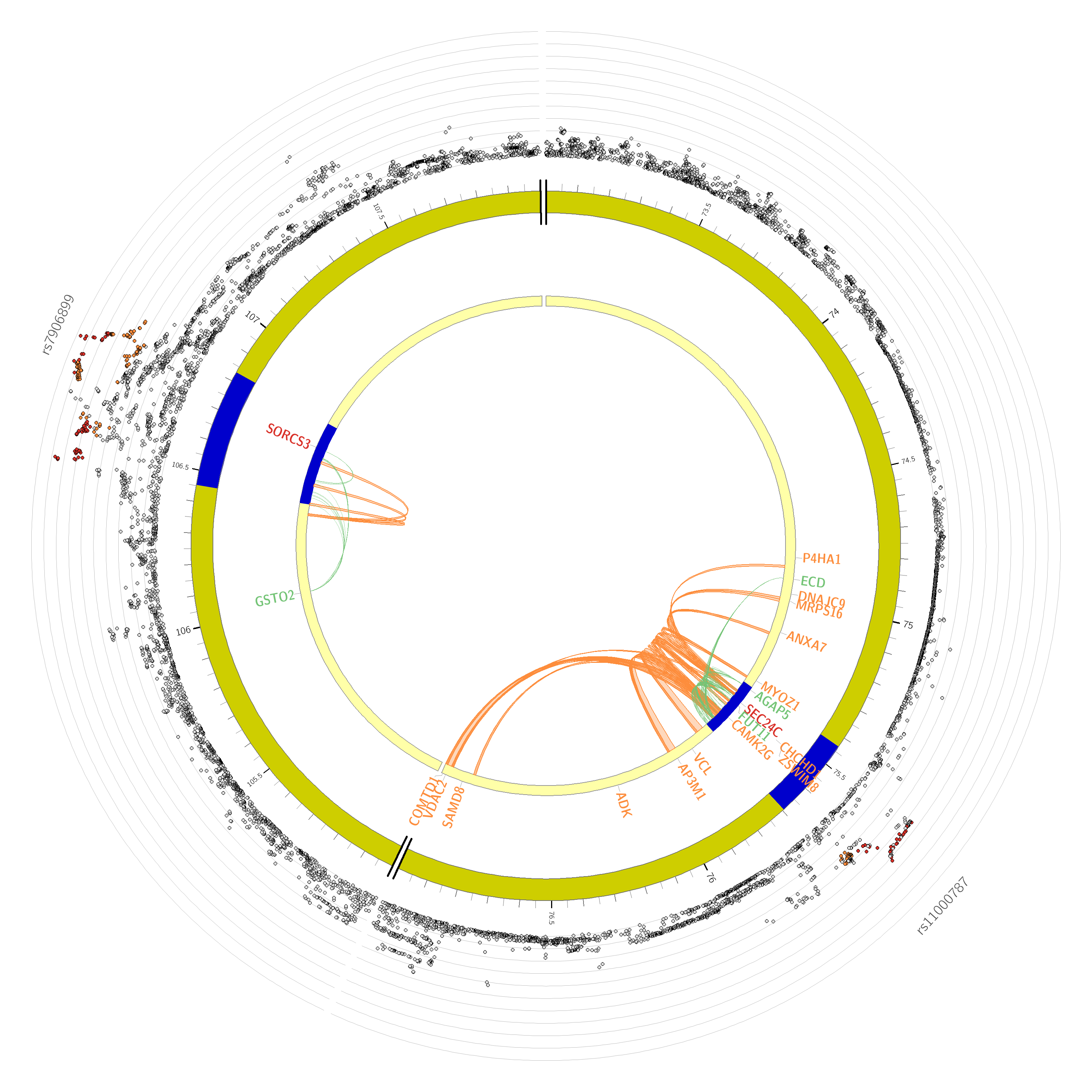

Chromosome 11

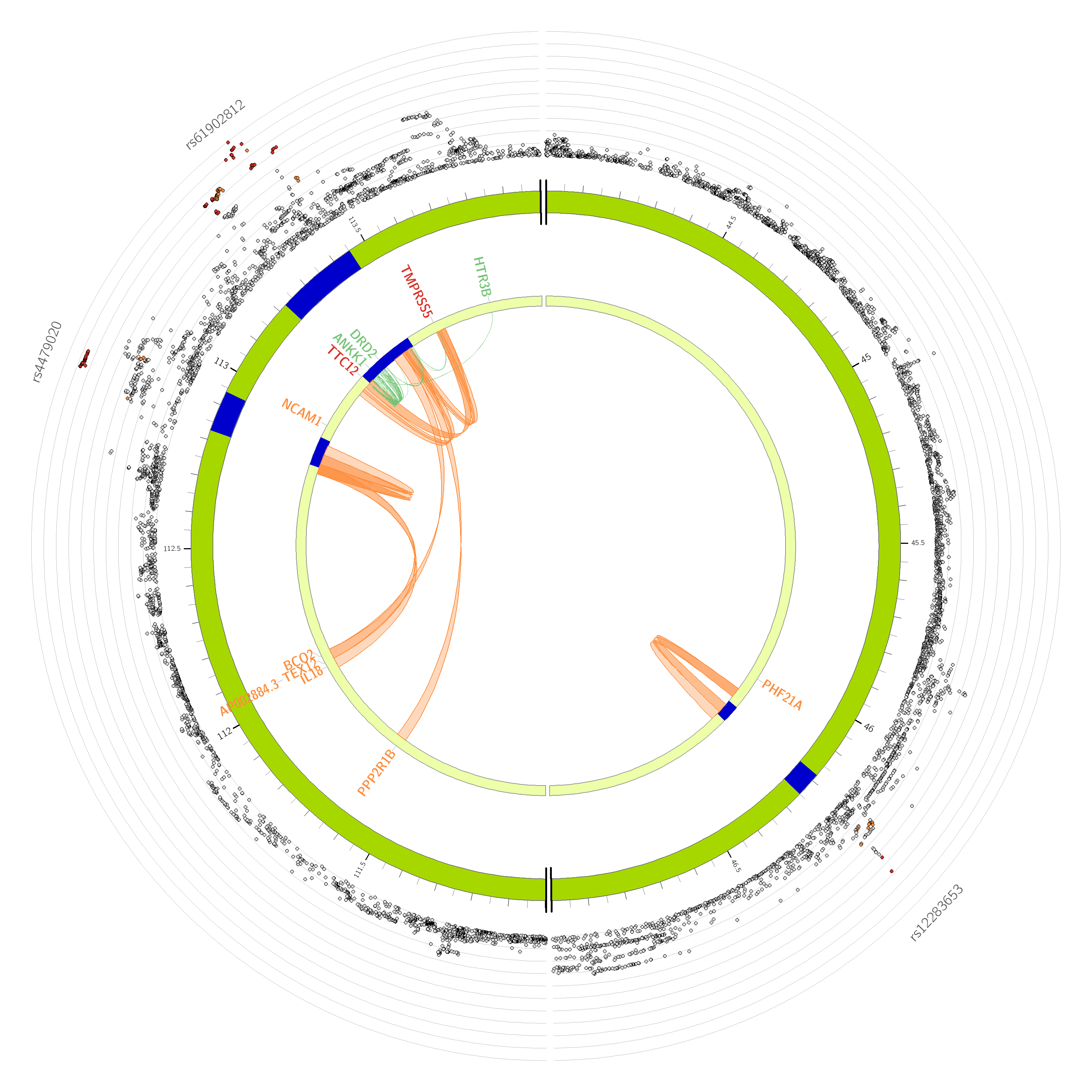

Chromosome 12

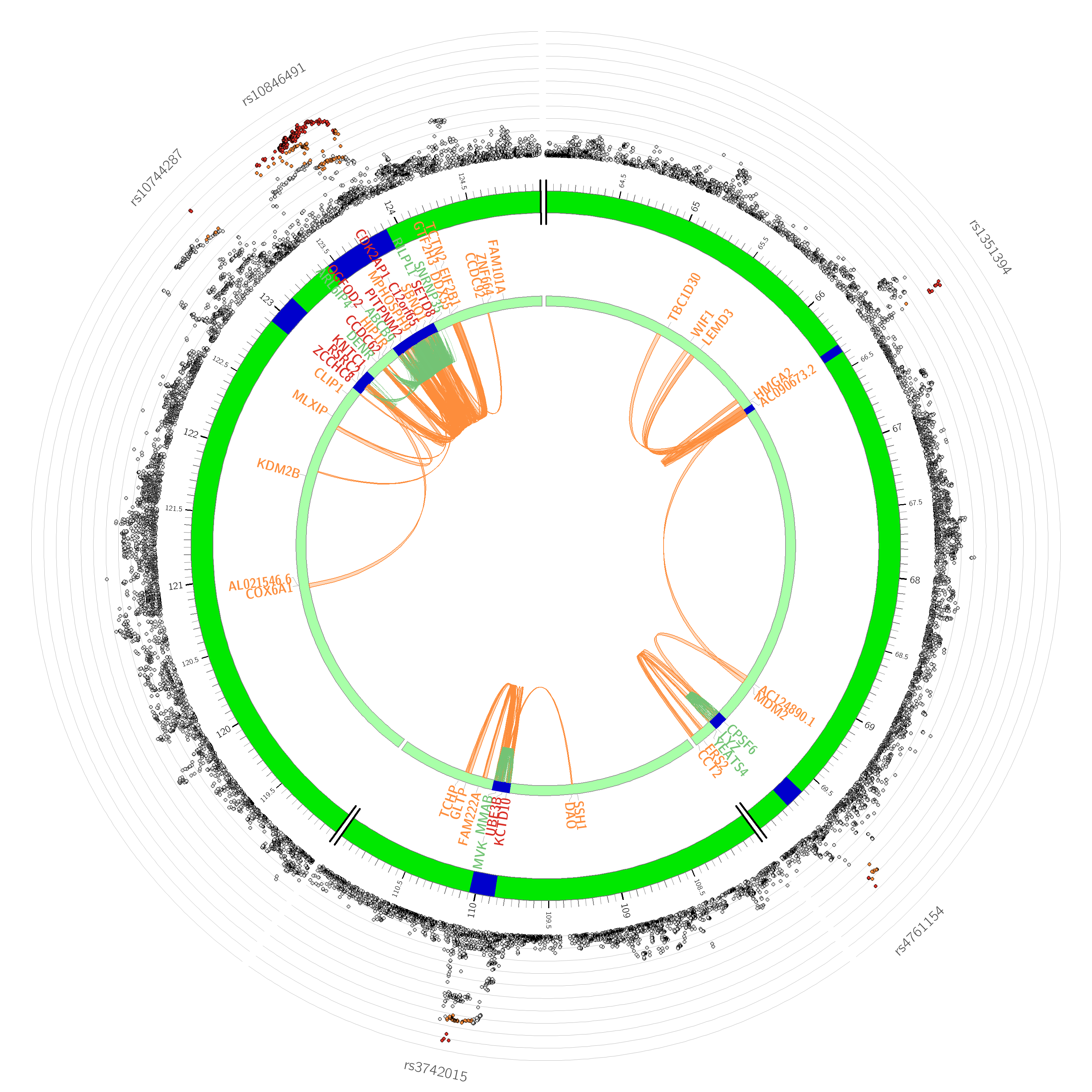

Chromosome 13

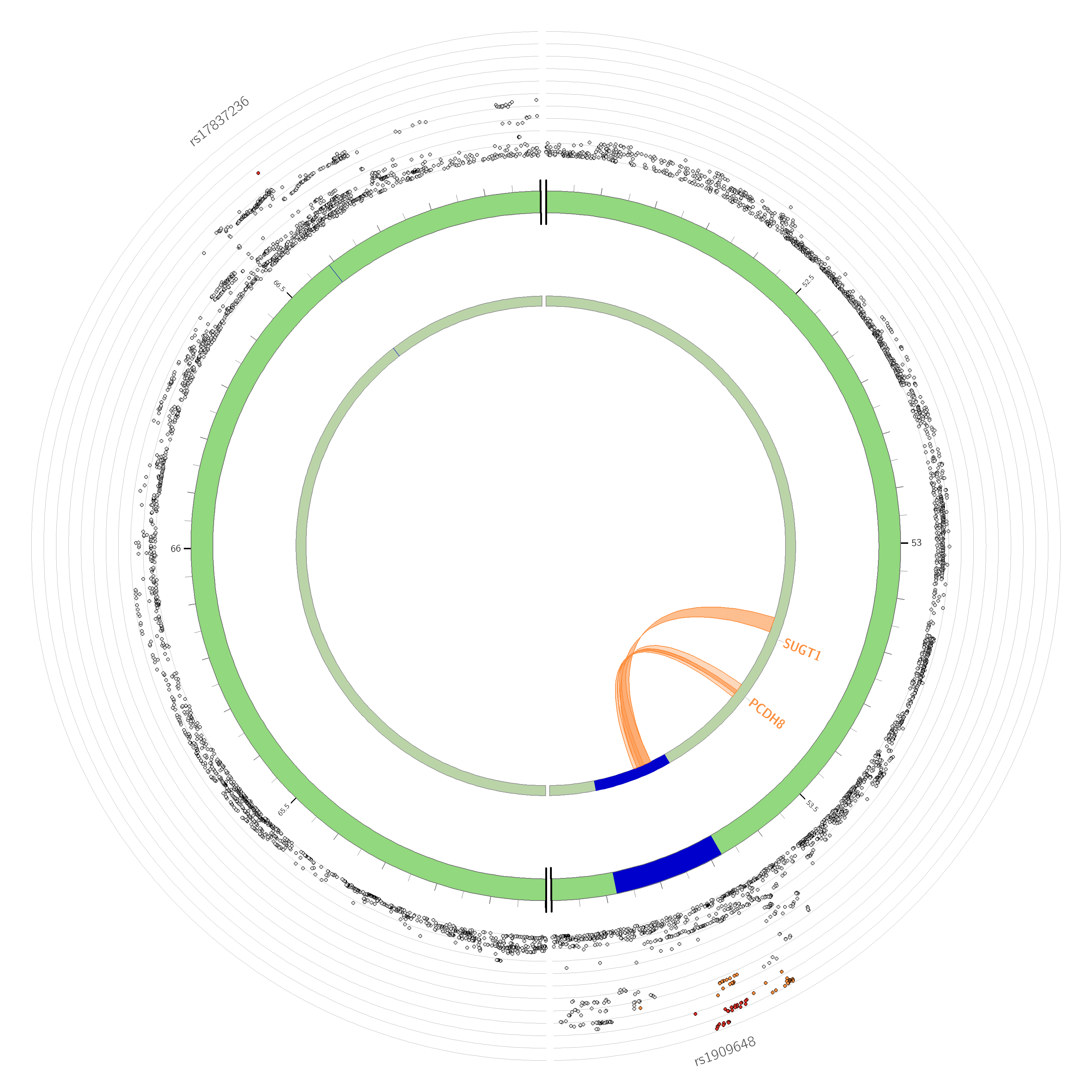

Chromosome 14

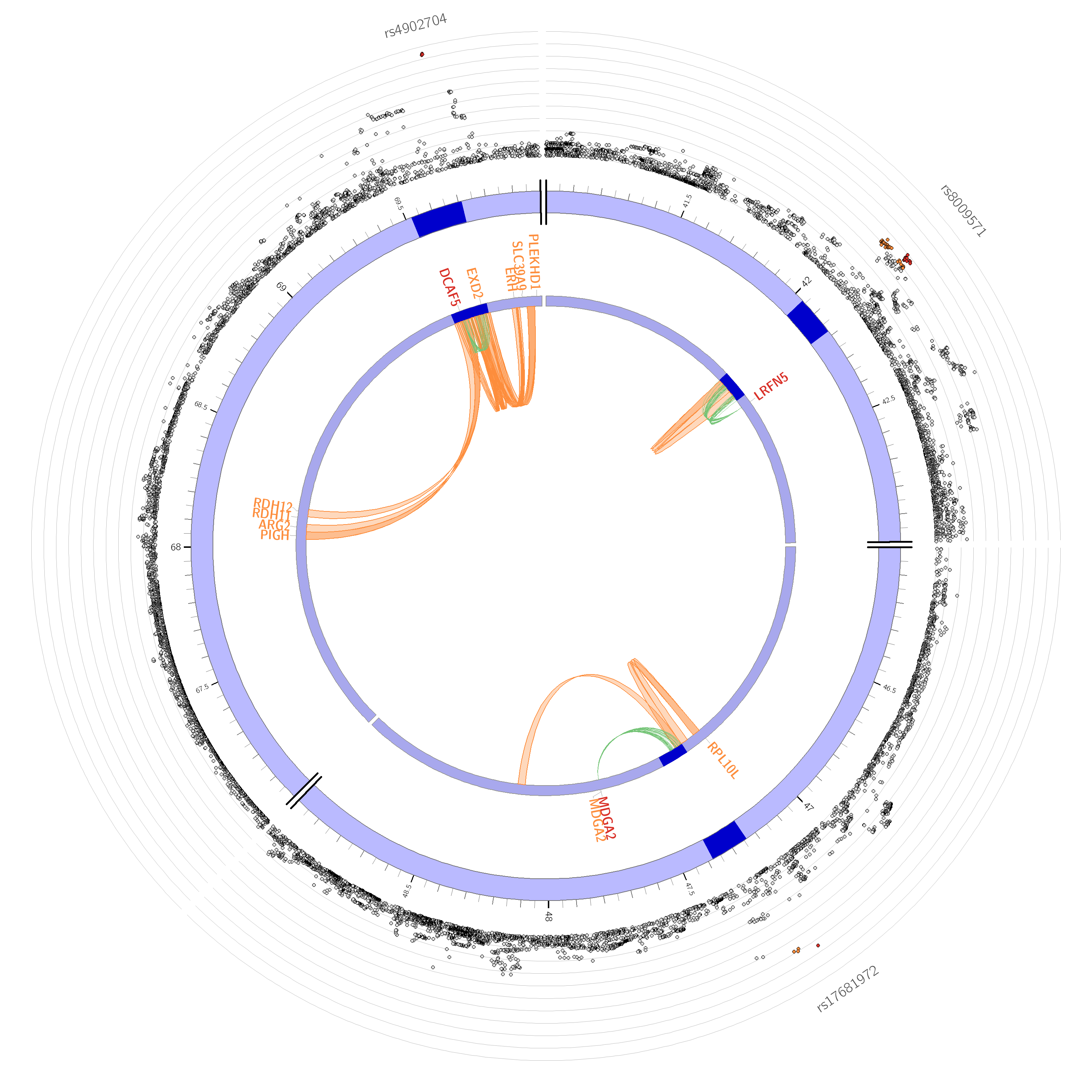

Chromosome 15

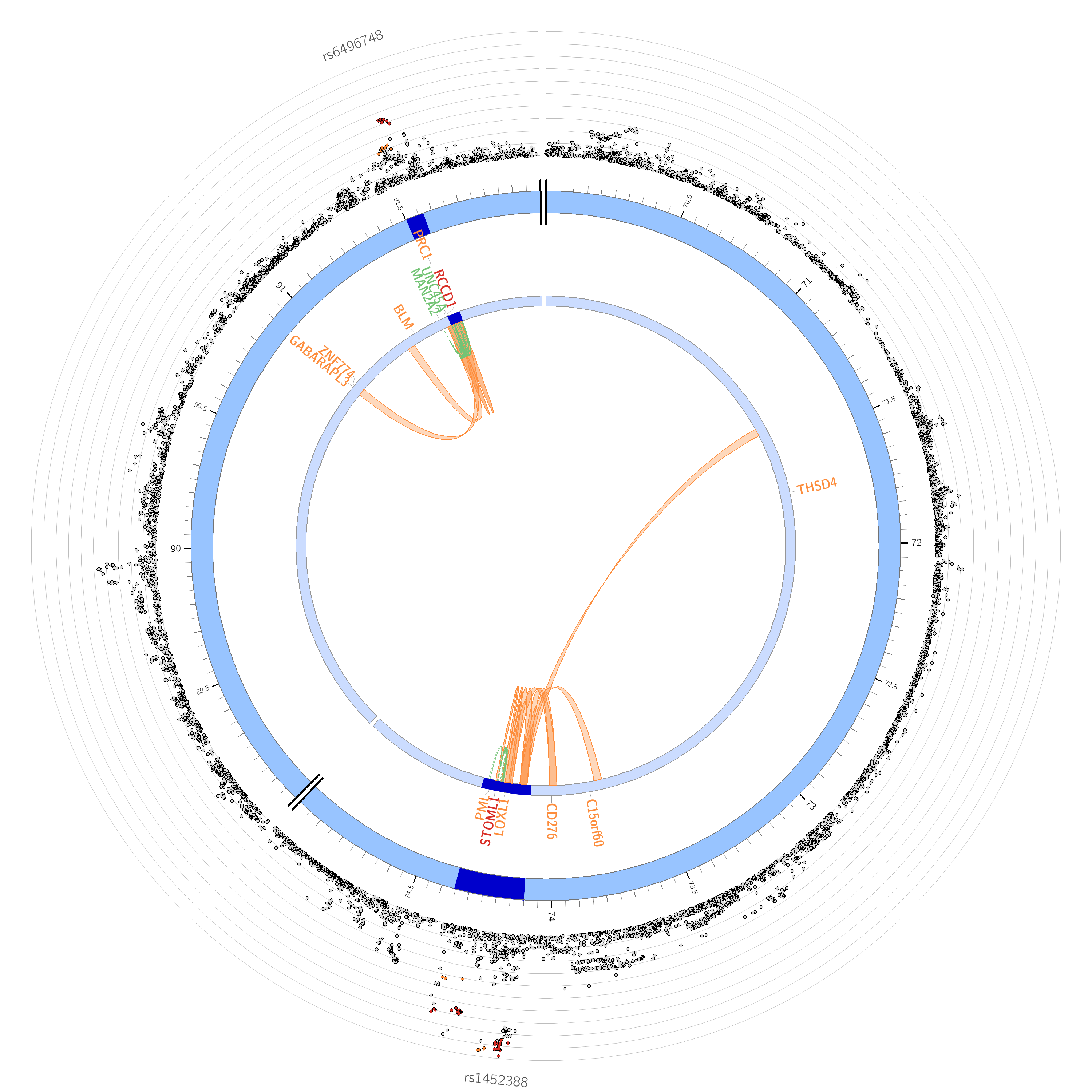

Chromosome 16

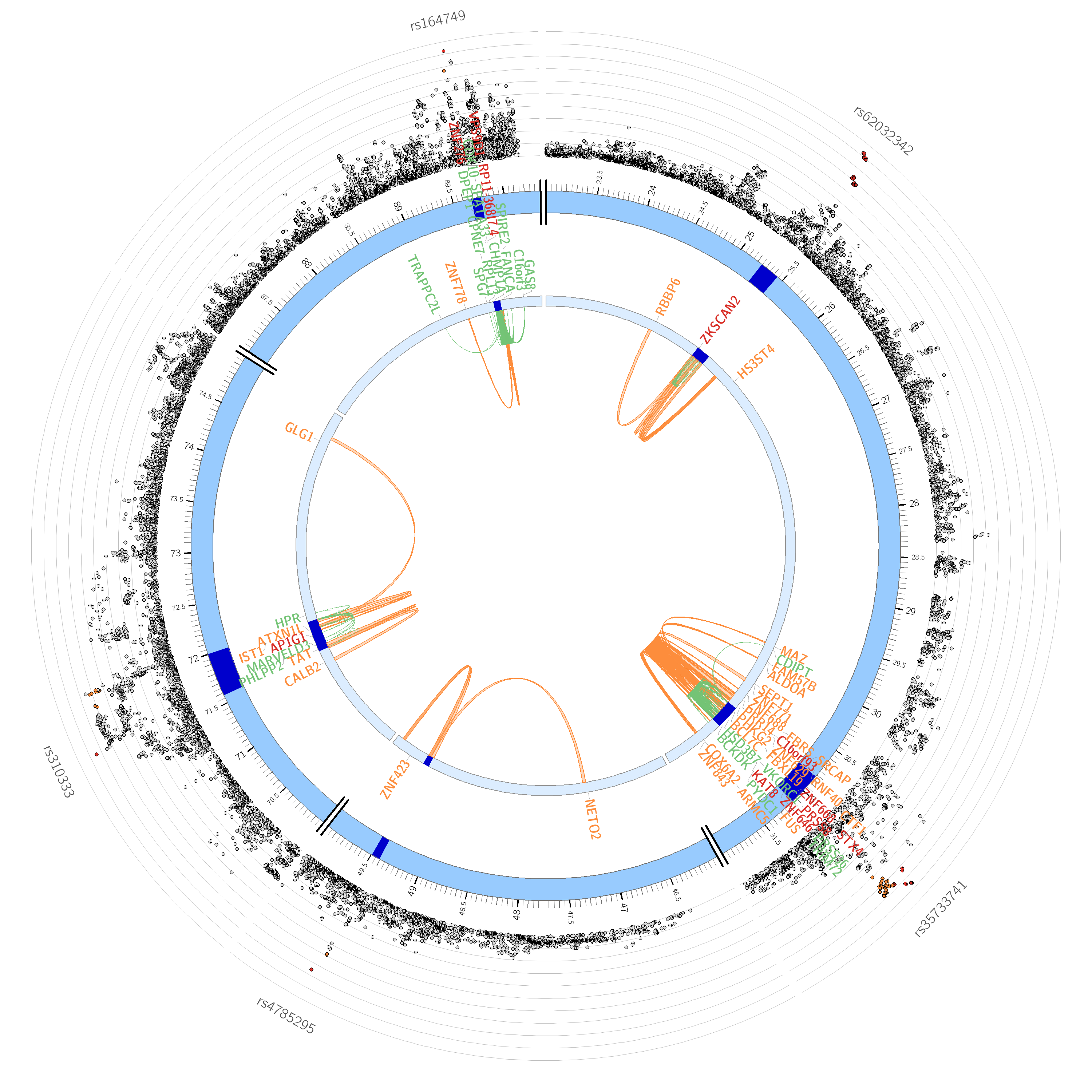

Chromosome 17

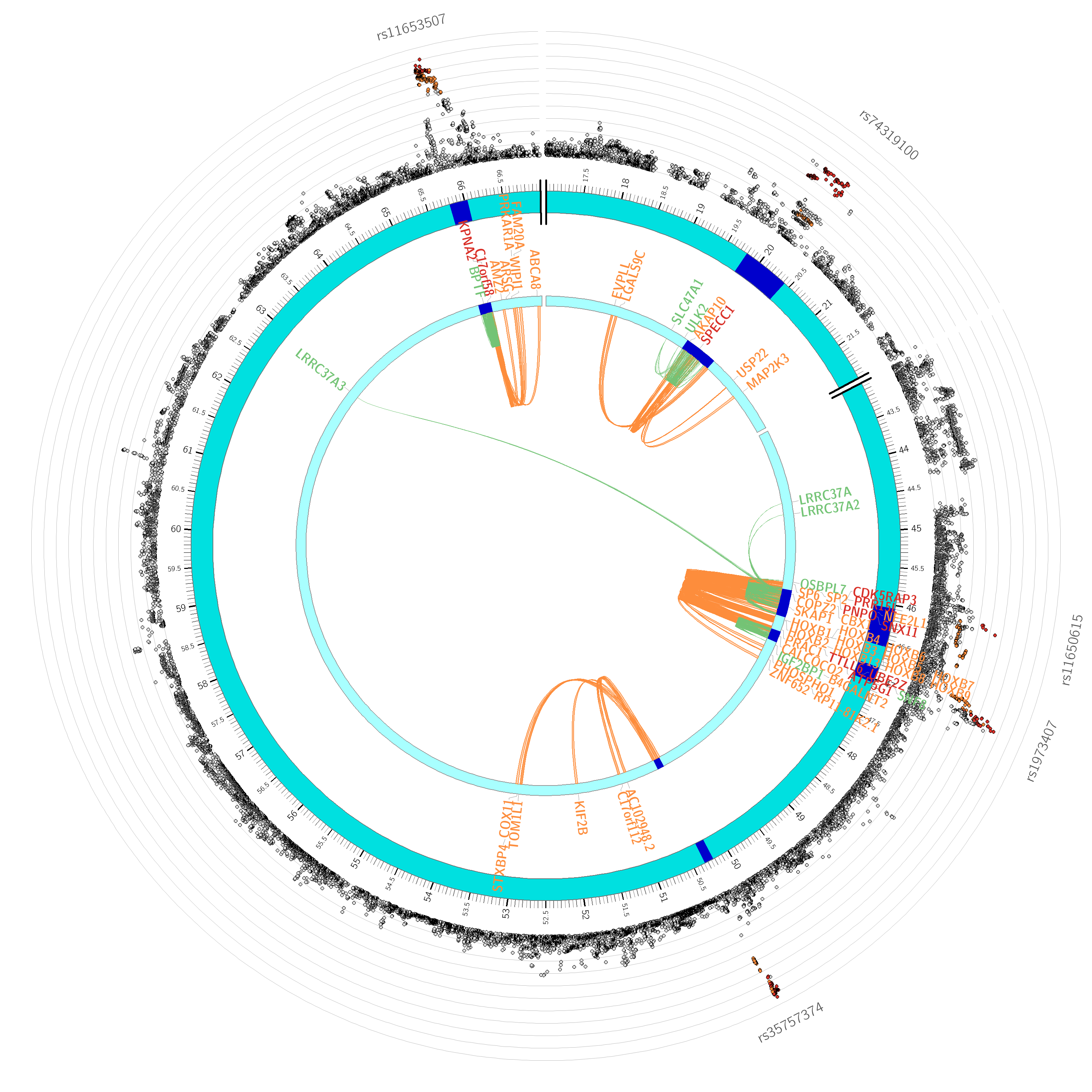

Chromosome 18

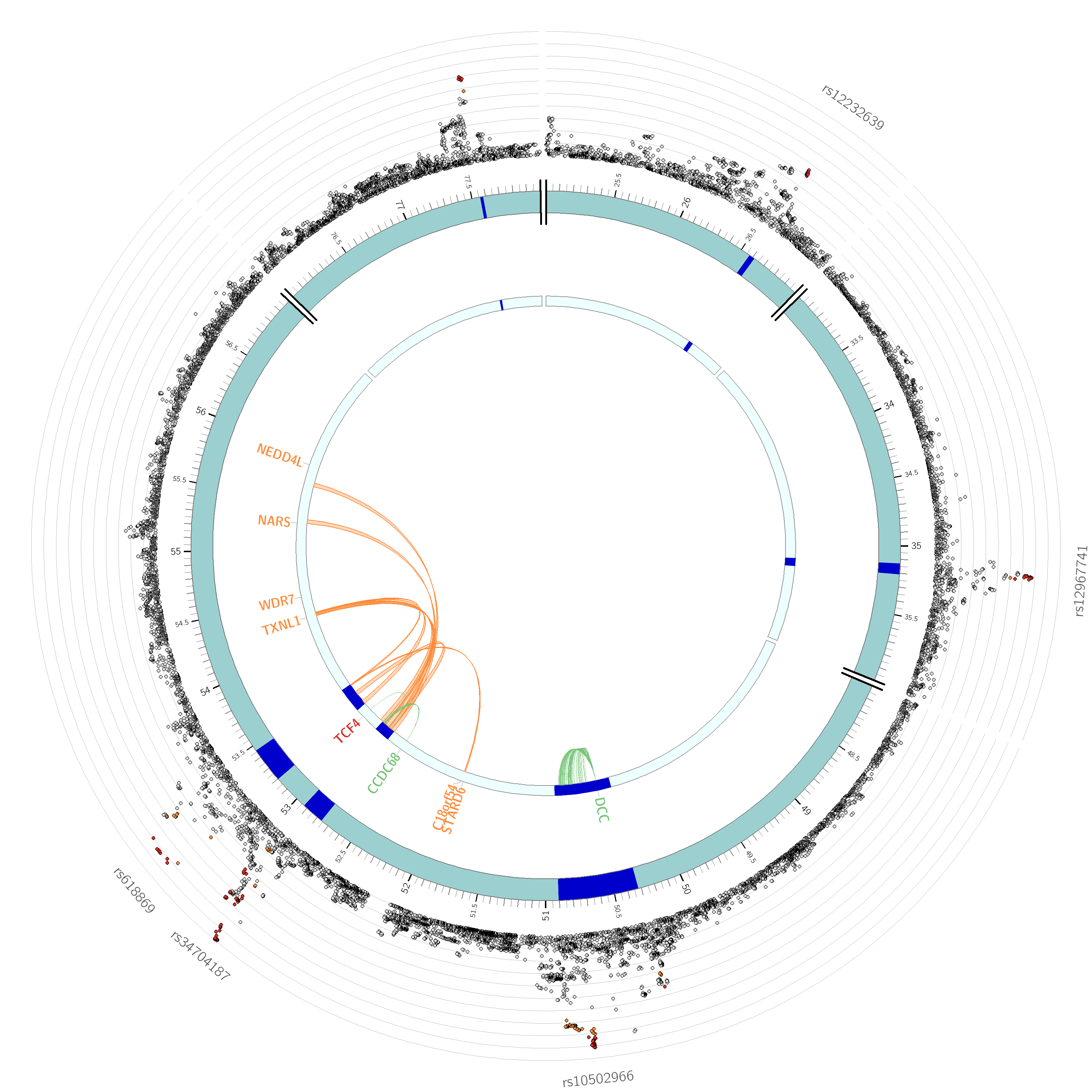

Only SNPs with *P* < 0.05 are displayed. SNPs in genomic risk loci are color-coded as a function of their maximum r^2^ to the one of the independent significant SNPs in the locus as follows: red (r^2^ > 0.8), orange (r^2^ > 0.6), green (r^2^ > 0.4) and blue (r^2^ > 0.2). SNPs that are not in LD with any of the independent significant SNPs (with r^2^ ≤ 0.2) are grey. The rsID of the top SNPs in each risk locus is displayed in the most outer layer. Y-axis ranges between 0 to the maximum -log10(P-value) of the SNPs. Genomic risk loci are highlighted in blue. Only mapped genes by either chromatin interaction and/or eQTLs are displayed. If the gene is mapped only by chromatin interactions or only by eQTLs, it is colored orange or green, respectively. When the gene is mapped by both, it is colored red.

Chromosome 19

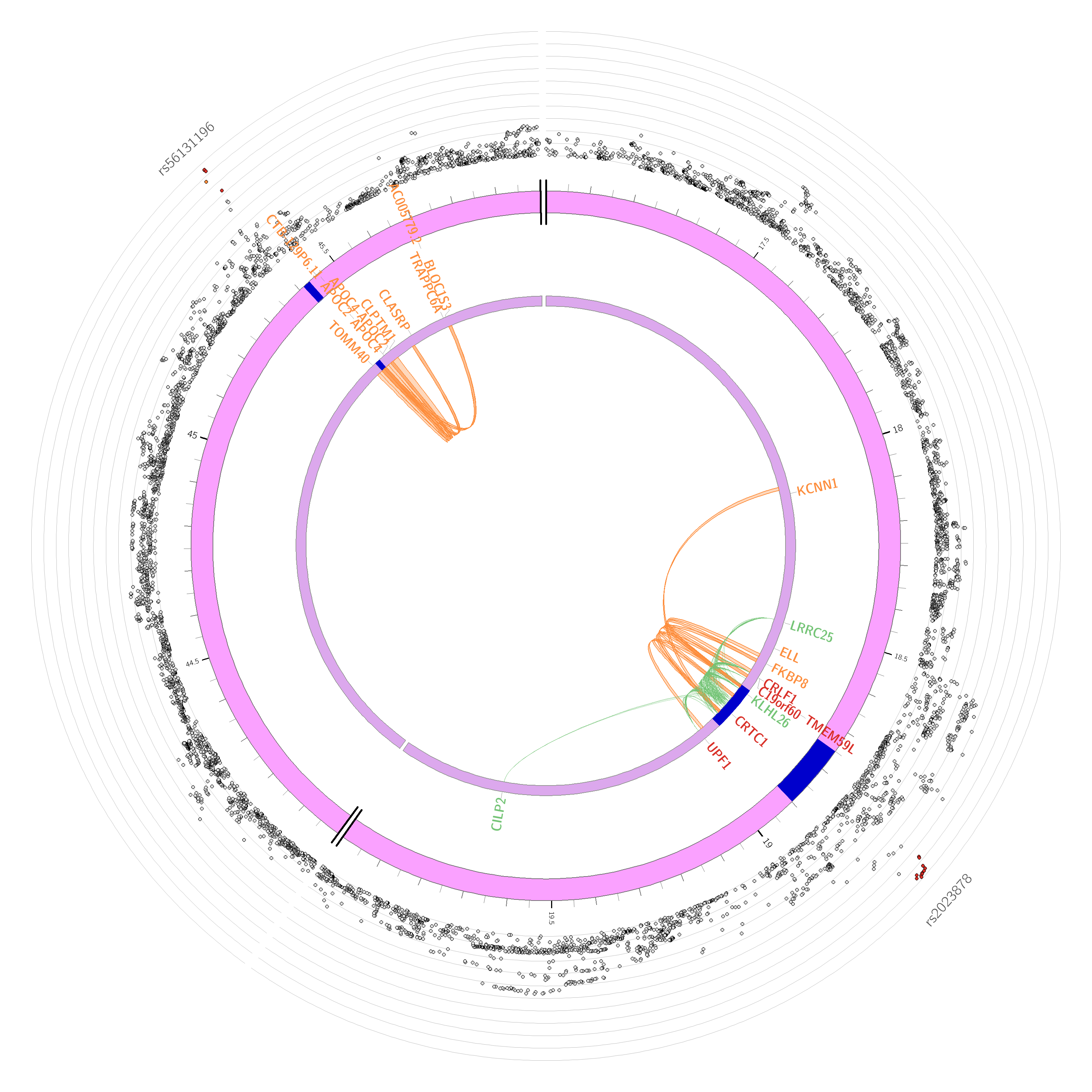

Chromosome 22

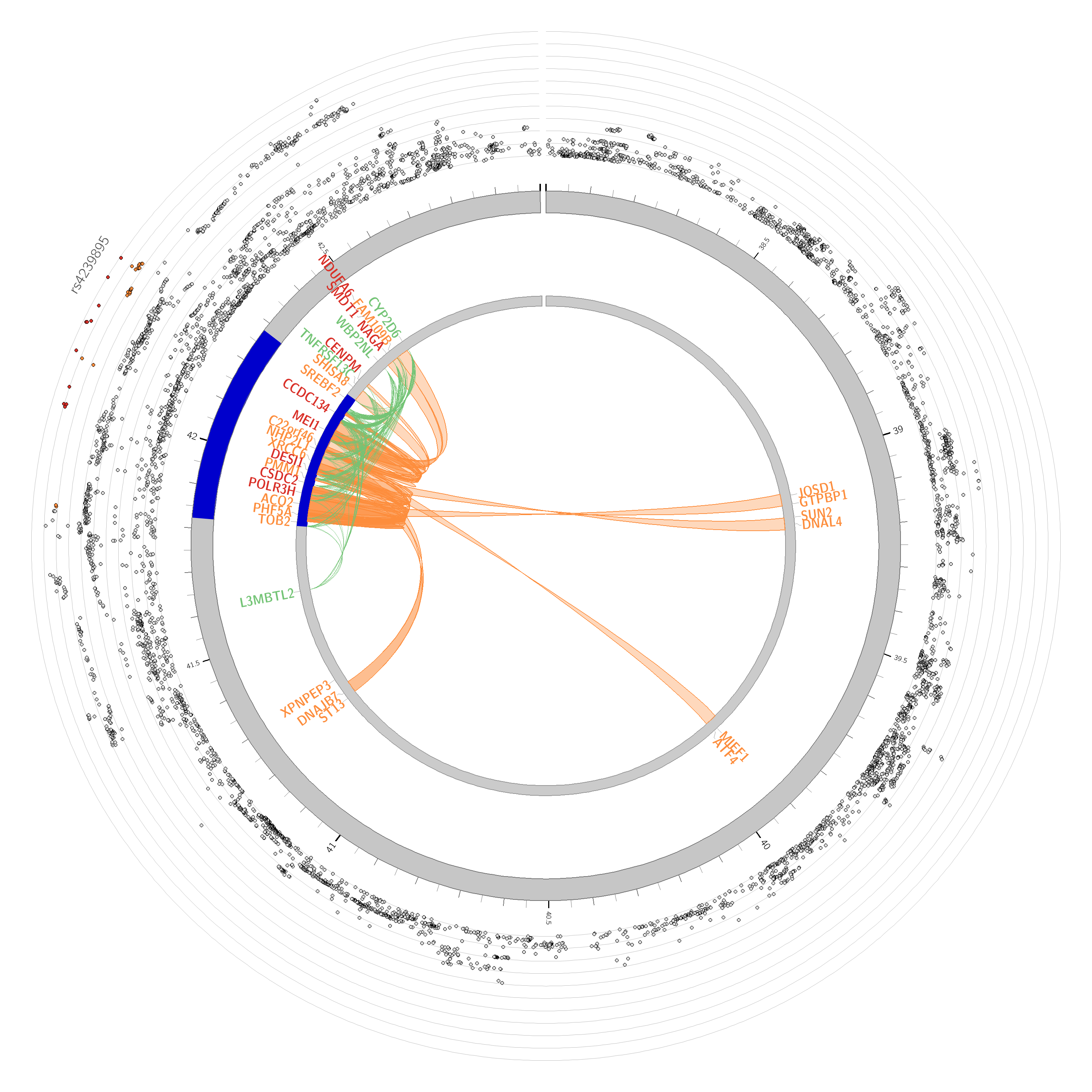

#### **Supplementary Figure 4. Gene-property analyses in BrainSpan.**

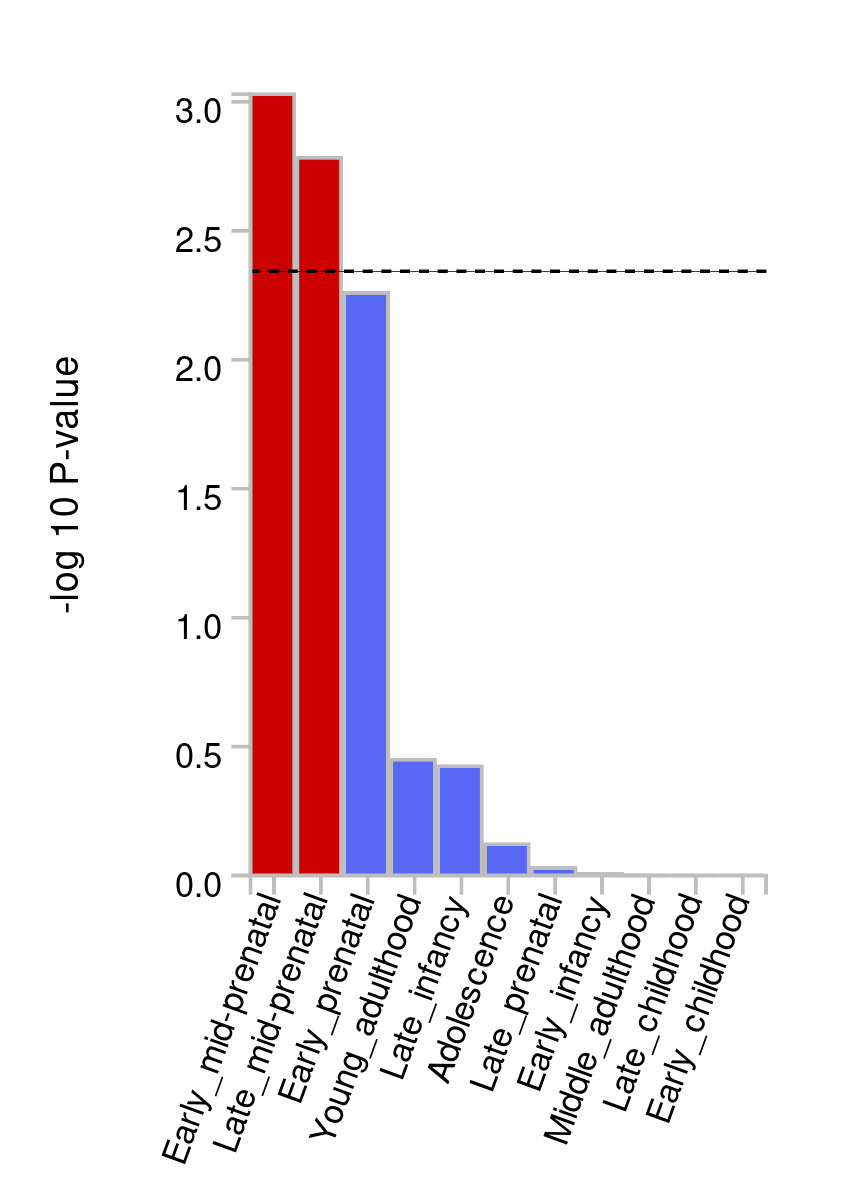

Red bars indicate developmental stages in which gene expression is significantly enriched. The dashed line is for a Bonferroni-adjusted *p* value.

#### **Supplementary Figure 5. Gene expression heatmap for genes identified in two independent TWAS.**

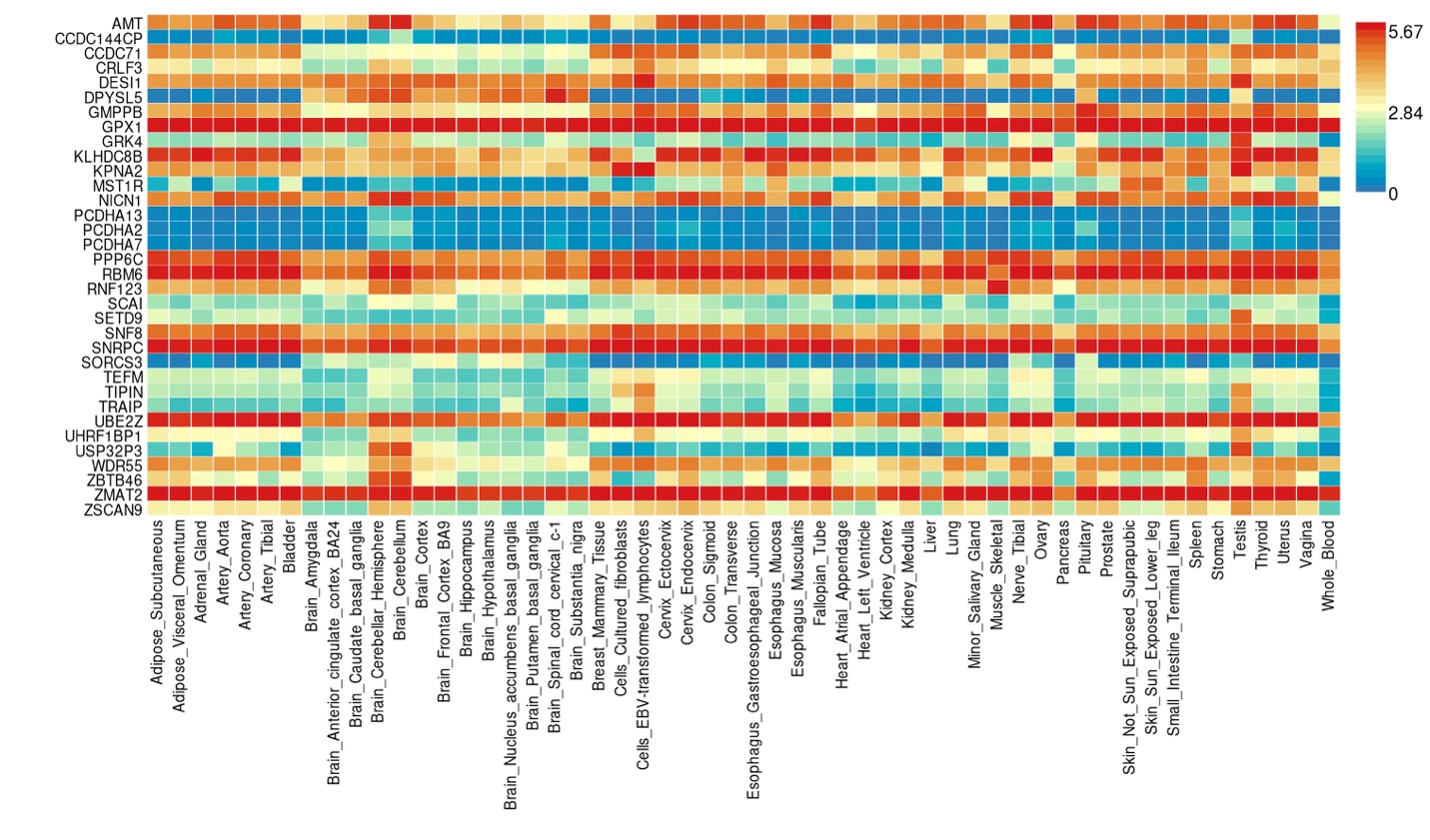

Gene expression values are based on average expression per label (log2 transformed) using Transcripts Per Million (TPM) normalization.

#### **Supplementary Figure 6. Bivariate MiXeR results examining polygenic overlap of the somatoform factor with psychopathology.**

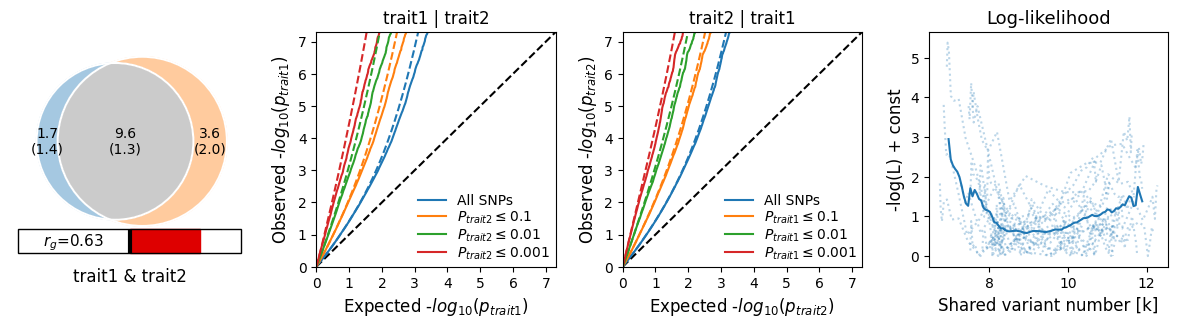

Somatoform & Internalizing

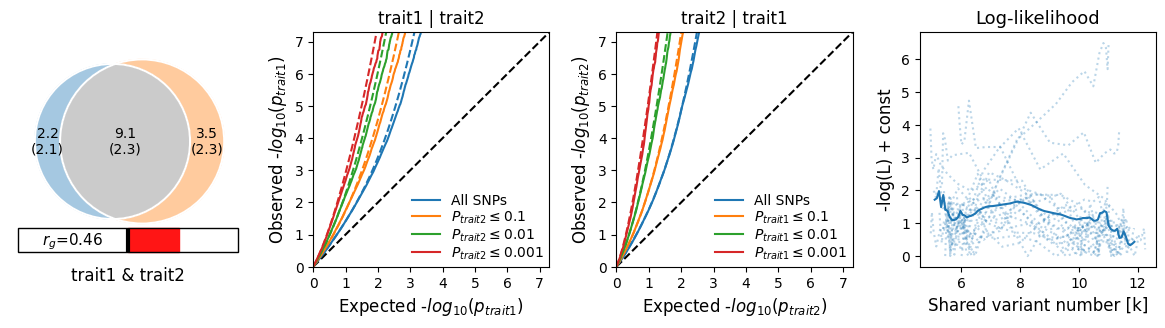

Somatoform & Externalizing

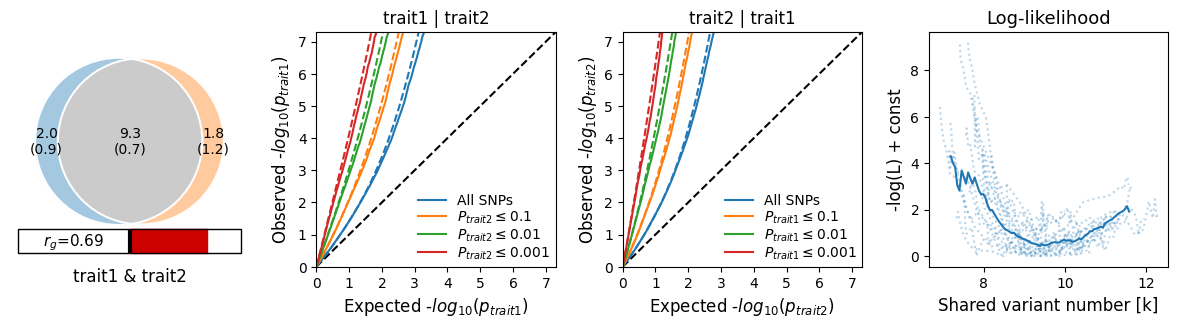

Somatoform & General Psychopathology

In all panels, trait 1 refers to the somatoform factor and trait 2 to the psychopathology factor.

#### **Supplementary Figure 7. Genetic correlations with publicly available GWAS.**

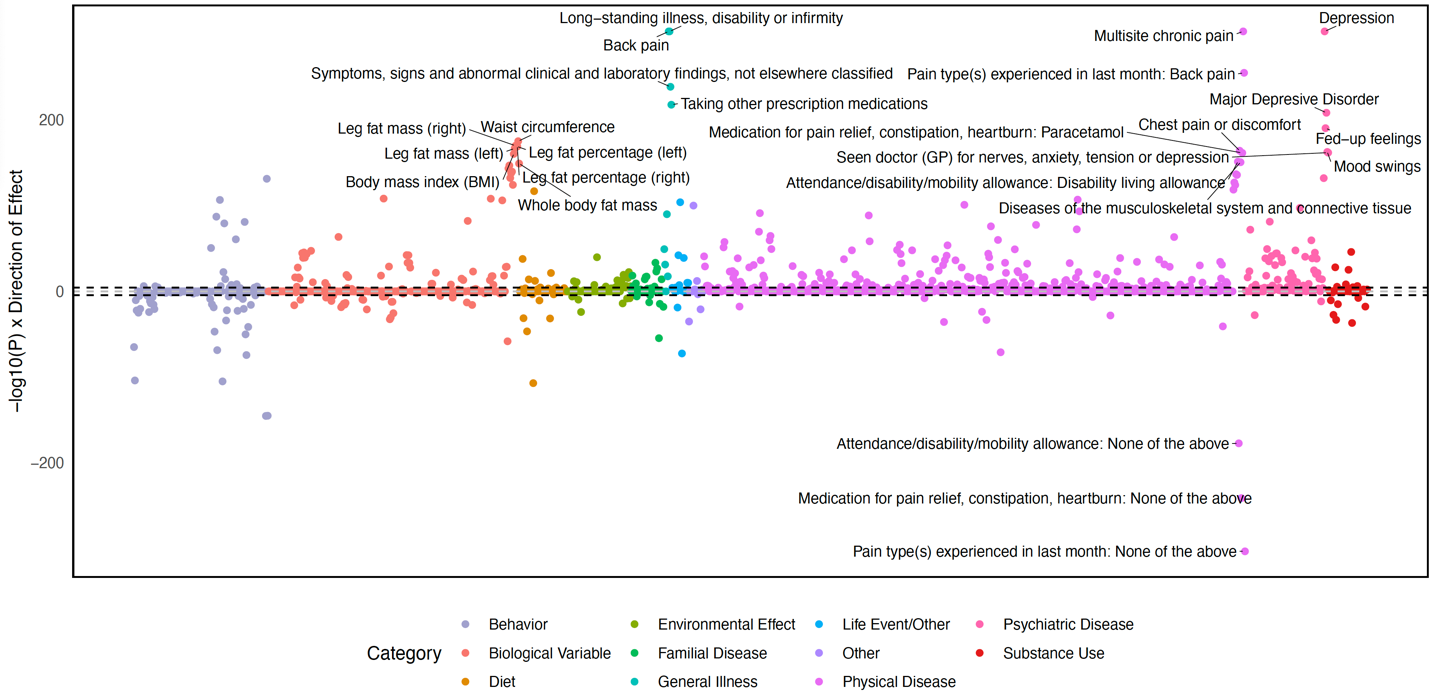

Dashed line indicates a Bonferroni-adjusted significant *p* value (0.05). Genetic correlations were calculated using the CTG-VL batch genetic correlations function.

#### **Supplementary Figure 8. LabWAS results in BioVU.**

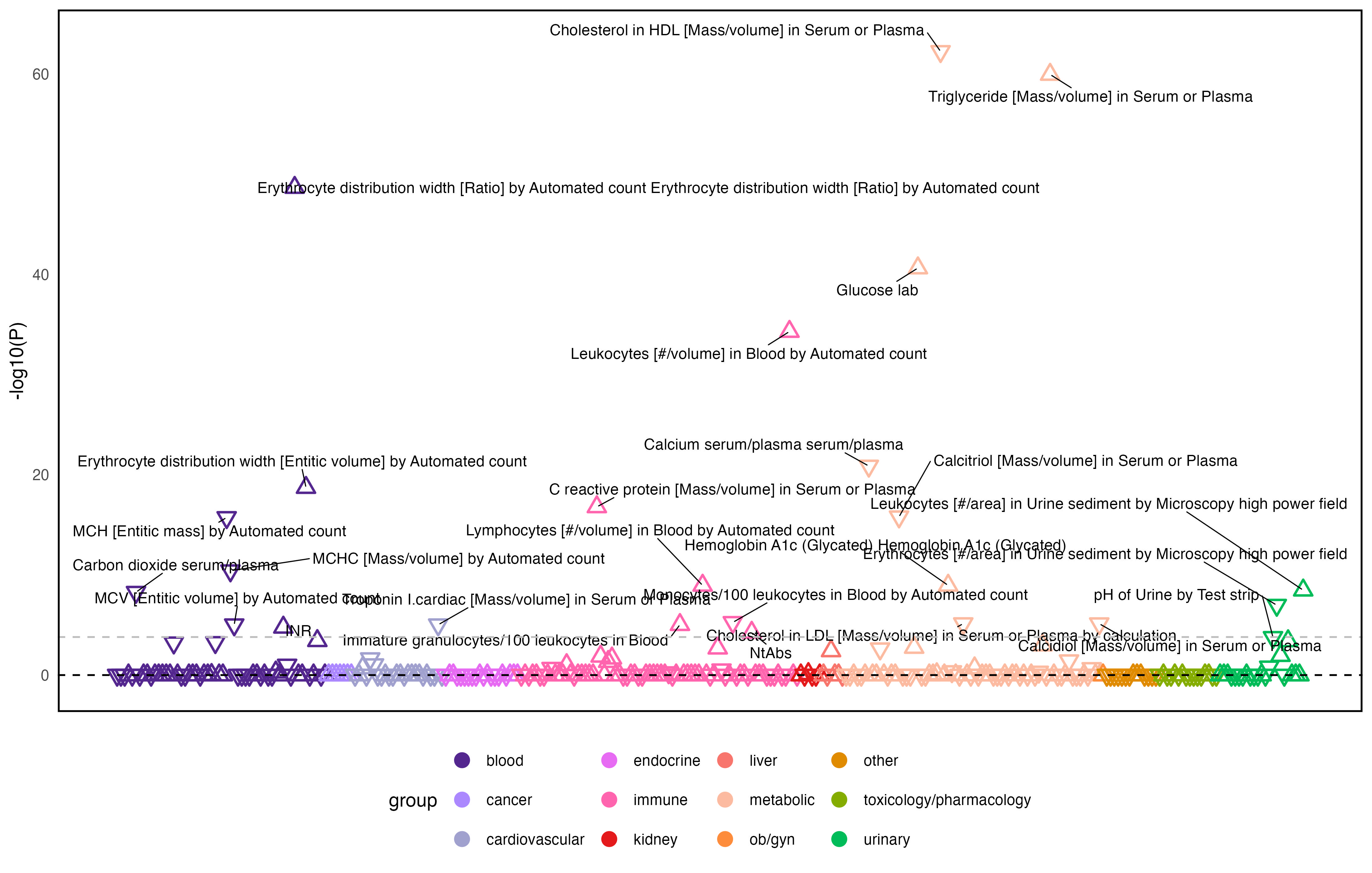

Dashed line indicates a Bonferroni-adjusted significant *p* value (0.05).

#### **Supplementary Figure 9. Sankey plot of drug repurposing results mapped to genome-wide significant genes.**

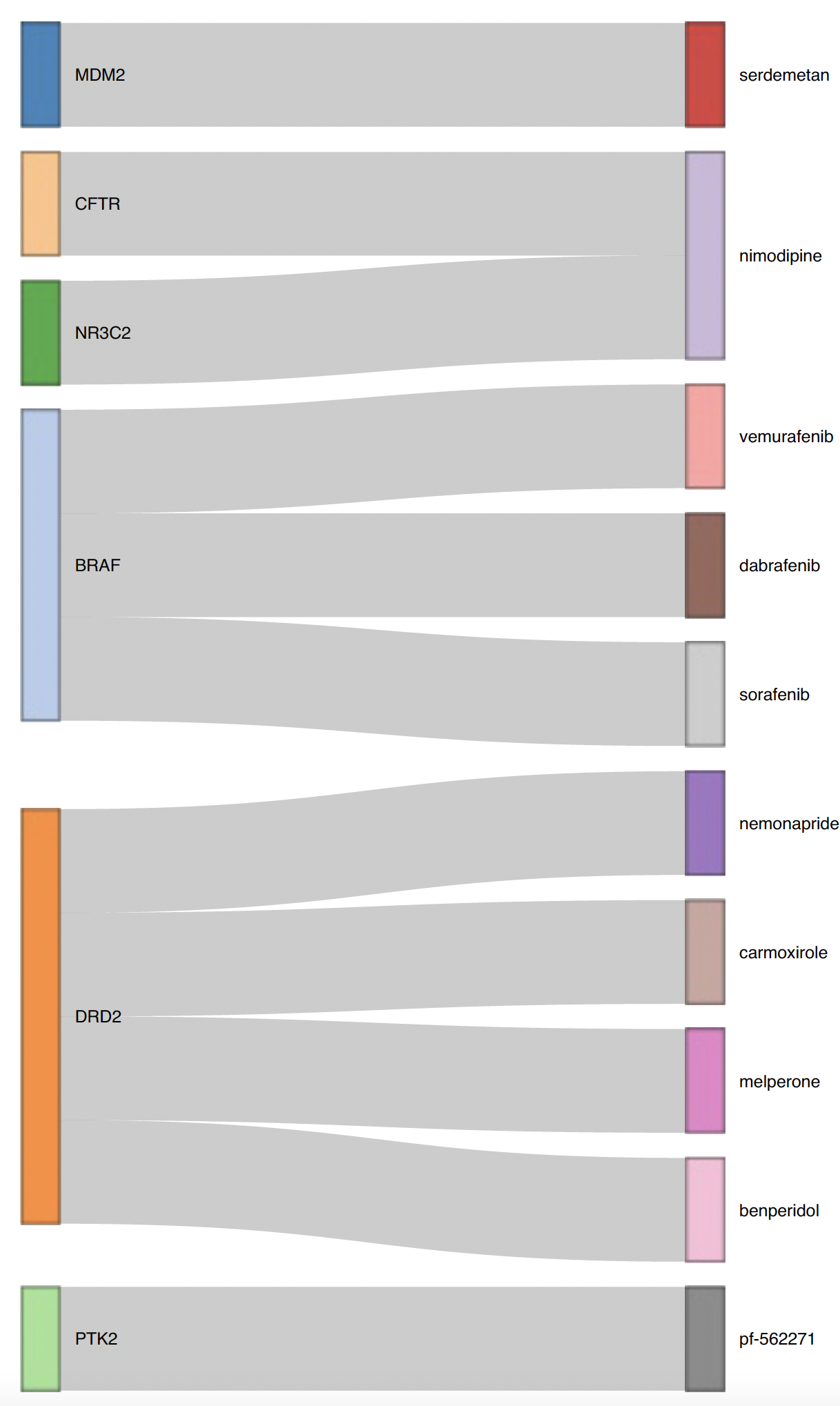

Genes were mapped based on position, eQTLs, and chromatin interactions using FUMA.

#### **Supplementary Figure 10. Mendelian randomization results examining causal effects of gut microbiota on the somatoform factor.**

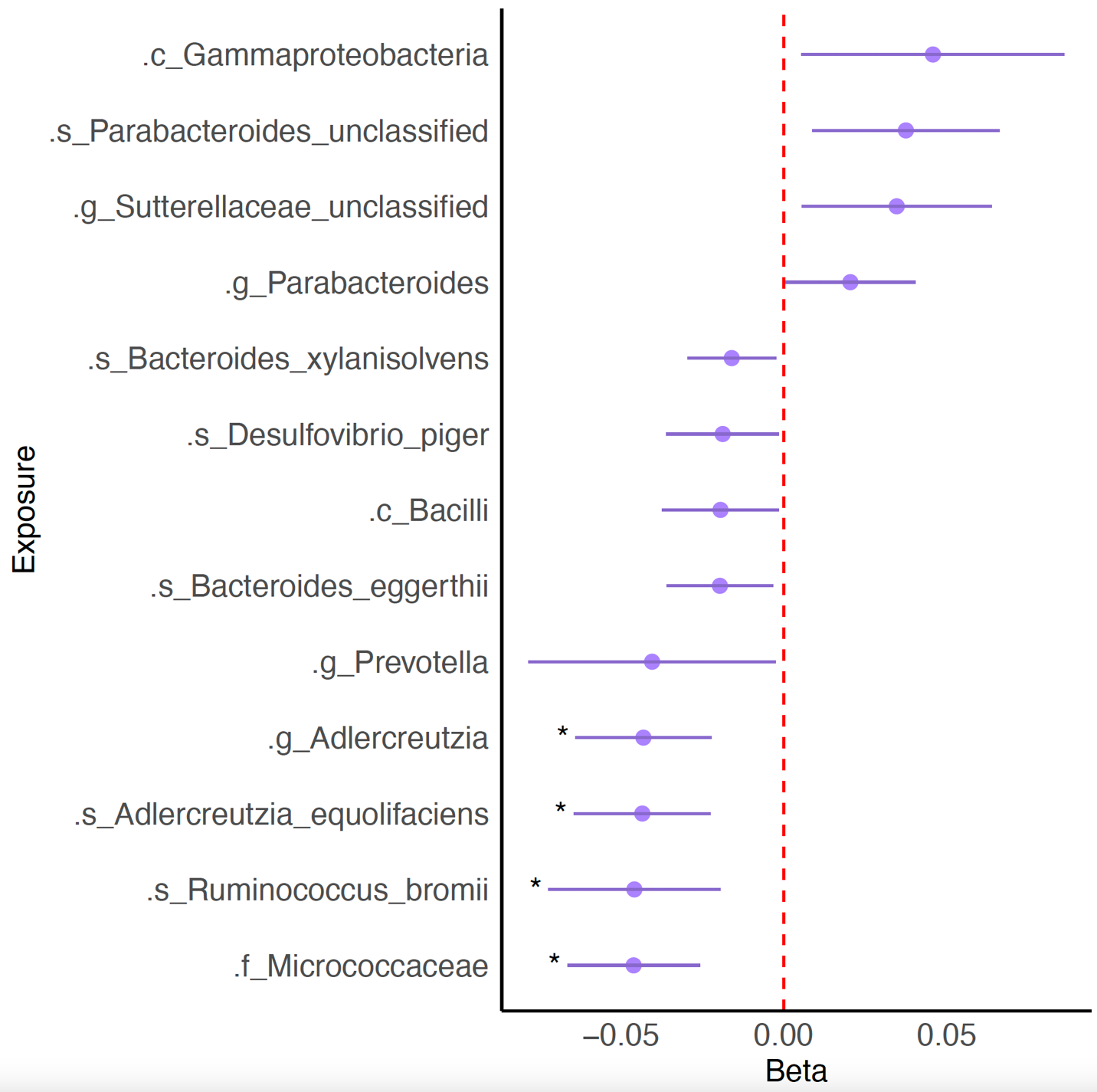

C = class, s = species, g = genus, f = family. Stars indicate significance after Bonferroni correction
